# Conditional normative modeling of whole-body FDG PET/CT: inter-organ relationships outperform reference-region normalization for label-free detection of lesion-bearing organs

**DOI:** 10.64898/2026.11.25.26369006

**Authors:** Chang-hyun Park, Hae-Jeong Park

## Abstract

**Purpose:** Reading whole-body 2-deoxy-2-[^18^F]fluoro-D-glucose (FDG) positron emission tomography/computed tomography (PET/CT) means judging each organ’s uptake against what is expected for that patient. Clinical criteria do this with a single reference region, the liver or the blood pool. We tested whether a normative model conditioned on all other organs detects lesion-bearing organs better than reference-region normalization, without lesion labels.

**Methods:** We analyzed the public autoPET FDG PET/CT dataset (1,014 studies; 896 patients with age and sex, 412 without malignant lesions). Models trained on control patients predicted the log maximum standardized uptake value (SUVmax) of each of 34 organs, segmented with TotalSegmentator on CT, from age, sex, and the median uptake of the other organs on eroded masks, without lesion labels, which served only as the reference standard. Out-of-fold deviation scores were compared with SUVmax, liver- and blood-pool-referenced, and age–sex normative scores for detecting organs with ≥ 2 ml of lesion (1,700 organs, 355 patients). Secondary analyses included false positives per patient, repeat scans, and residual soft-tissue regions.

**Results:** Other organs explained a median 67% of the between-patient variance in uptake, against 34% for global uptake with age and sex. The conditional linear model outperformed liver referencing: the within-patient area under the receiver operating characteristic curve (AUC) was 0.916 vs 0.902 (difference 0.014, 95% confidence interval [CI] 0.007–0.021), the false-positive rate at 90% sensitivity fell from 0.33 to 0.27, and at 80% sensitivity 2.9 instead of 4.3 lesion-free organs per patient were flagged. A global-uptake reference gained little (AUC 0.907). The advantage was robust to analysis choices, including a peak SUV target, and to lesion-cleaned inputs, and leveled off at about 200 training controls. Gradient boosting did not improve on the linear model, whereas modeling patient-specific variance made calibration uniform across patients. Conditional scores changed organ by organ between repeat scans, and the gain extended to mediastinal but not abdominal or pelvic soft tissue.

*Conclusion:* Conditioning an organ’s expected uptake on the patient’s other organs generalizes reference-region normalization and reduces false positives in label-free detection of abnormal organ uptake, using oncology controls rather than healthy volunteers. Organ-level conditional deviations provide a calibrated, patient-specific reference on which voxel-level localization within organs can build.

## 1. Introduction

Whole-body 2-deoxy-2-[^18^F]fluoro-D-glucose (FDG) positron emission tomography/computed tomography (PET/CT) measures glucose metabolism in every organ in one examination [1]. Whether an organ’s uptake is abnormal depends on the patient: FDG uptake rises with inflammation, infection, and normal physiological variation as well as with tumor [2], and it shifts with blood glucose, uptake time, and body habitus [3, 4]. Readers therefore judge uptake against a reference within the same scan.

Clinical criteria formalize this with one reference region: Deauville scoring for lymphoma compares lesion uptake with the liver and the mediastinal blood pool [5], and the PET Response Criteria in Solid Tumors (PERCIST) define measurable disease relative to liver uptake [6]. Such referencing is a conditional normalization on a single organ with a fixed slope of one, and it ignores what the rest of the body says about the expected uptake of the organ being read.

Normative modeling offers a more general approach: it estimates the distribution of a measurement in a reference population and scores each individual as a deviation from it [7, 8]. Most normative models condition only on covariates such as age and sex (marginal models). A recent model of cortical thickness instead reconstructed each cortical location from the subject’s other locations and found deviations more closely tied to diagnosis than those of marginal models [9]. Whole-body PET is a natural setting for this conditional approach, because organ uptake values are correlated within a person [10] and inter-organ metabolic networks have been described at the group level [11–14]. Individual-level work has so far compared a patient’s inter-organ network with a healthy or demographically matched reference [10, 13–15] or predicted a voxel-wise "normal twin" PET from CT and acquisition attributes [16]. None of these studies scores each organ against the expectation set by the patient’s own uptake in other organs, and none has been compared with the reference-region normalization used in clinical criteria.

We chose organs, rather than voxels, as the unit of this conditional reference. In the brain, voxels or cortical surface vertices can be brought into correspondence across subjects by registration, which is what makes location-wise normative models possible [8, 9]. Body habitus, bowel content, bladder filling, and breathing make such correspondence unreliable for the trunk, whereas automated CT segmentation gives every patient the same set of organs, and clinical reading criteria are themselves expressed per organ or reference region [5, 6].

Two further problems stand in the way of such a reference: healthy whole-body PET scans are scarce because of radiation exposure, and the large public datasets come from oncology patients [17], and deviation scores must be calibrated, because a score that flags variable organs too often adds false positives to the reader’s workload.

We asked three questions using the public autoPET FDG dataset. First, how much of an organ’s uptake is explained by the uptake of other organs, beyond age, sex, and global uptake? Second, does a normative model conditioned on other organs detect lesion-bearing organs better than maximum standardized uptake value (SUVmax), liver- or blood-pool-referenced normalization, and a marginal normative model? Third, do non-linear models, or a variance model of aleatoric variability, the patient-specific spread of uptake around the expected value,, improve detection or calibration?

## 2. Materials and Methods

### 2.1 Data

We used the public autoPET FDG PET/CT dataset (FDG-PET-CT-Lesions) [17, 18] in its openly downloadable NIfTI release (University of Tübingen data repository); the original DICOM files are hosted by The Cancer Imaging Archive (TCIA). It contains 1,014 whole-body studies from 900 patients examined at a single center, comprising patients with malignant lymphoma, melanoma, or non-small cell lung cancer and patients without malignant FDG-avid lesions, who served as controls and are not healthy volunteers. Manual segmentations of FDG-avid tumor lesions were created and released by the dataset curators [17] and served, without modification, as the lesion reference. We used the standardized uptake value (SUV) images and the CT resampled to the PET grid (2.04 × 2.04 × 3 mm) that are distributed with the dataset, so CT-derived masks apply to PET voxels without further resampling. We analyzed the first study of each patient with recorded age and sex.

### 2.2 Organ segmentation and features

TotalSegmentator v2 [19] segmented 117 structures on the CT at 1.5 mm resolution (body crop and split inference). Structures were merged into 41 organ groups (Supplementary Table S1); for example, all thoracic vertebrae formed one group, all ribs another, and the two kidneys were kept separate. For each group we computed SUV statistics on three masks: the full mask; the mask eroded by 4 mm to limit partial-volume and misregistration effects; and a "clean" mask, the eroded mask minus lesions dilated by 4 mm. Deviation scores used only the full and eroded masks; the clean mask, which requires the lesion labels, served for descriptive analyses and a sensitivity analysis. Organs cut by the scan range were set to missing. Respiratory PET–CT misregistration was indexed by the mean SUV of the upper 15 mm of the liver mask divided by that of the eroded liver; values below 0.7 indicate lung included in the liver mask [20].

Three organs were excluded a priori: the urinary bladder, whose uptake reflects urinary excretion of FDG rather than tissue metabolism; the prostate, which is present only in men; and the gallbladder, which is often surgically removed. Organs with a usable eroded value (not truncated, at least 20 voxels) in fewer than 90% of controls were also excluded. Missing predictor values of the remaining organs were imputed with the median of the training controls within each cross-validation fold.

### 2.3 Inter-organ relationships

In controls we computed Spearman and partial correlations (age and sex removed) of the log eroded median SUV, which equals the clean value in controls. For each organ we predicted its log eroded median SUV with ridge regression from four predictor sets: age and sex (M0); M0 plus global uptake, the mean of the other organs (M1); M0 plus every other organ (M2); and M0 plus the other organs outside the target’s tissue compartment (M2c). Compartments were bone (12 skeletal groups), blood pool and vessels (5), skeletal muscle (3), lung and airway (3), gastrointestinal tract (5), and kidney (2); two of the skeletal groups were later excluded (Section 3.1), and the liver, spleen, heart, thyroid, and the remaining organs belonged to no compartment (Supplementary Table S1). Performance was the out-of-fold coefficient of determination (*R*^2^) from 5-fold cross-validation repeated 20 times. *R*^2^ was computed within each organ and therefore counts only between-patient variance. An *R*^2^ pooled over all organ–patient cells would also credit the differences between organs (Section 3.2).

All models were fitted with scikit-learn (Section 2.12). Ridge regression, linear regression with an L2 penalty on the coefficients, was the primary model. The predictors, the uptake of the other organs, are many and strongly correlated, whereas the controls number a few hundred; the penalty stabilizes the weights of correlated predictors, and the model remains linear, so that its coefficients generalize the single fixed weight of reference-region normalization. The penalty was selected from 13 log-spaced values between 10^−3^ and 10^3^ by efficient leave-one-out cross-validation within the training data (RidgeCV); for the deviation scores (Section 2.4), predictors were first standardized on the training data. Gradient boosting of histogram-based regression trees (HistGradientBoostingRegressor) served as the non-linear alternative, because it captures non-linear relationships and interactions between organs without specifying them in advance; it was used only for the deviation scores.

### 2.4 Deviation scores

The target was each organ’s log SUVmax on the full mask, which includes any lesion voxels and, unlike the eroded median of Section 2.3, responds to focal lesions; every method turns it into a standardized deviation *z*, where a high *z* means more suspicious uptake. The referenced methods are special cases of the conditional model: liver referencing is a regression on one organ with its slope fixed at one; Figure 1 shows both scores in one patient.

1. **SUVmax:** log SUVmax standardized per organ.
2. **Liver-referenced:** log(SUVmax / liver mean), with the liver itself referenced to the aortic blood pool. Reference uptake was the mean SUV of the eroded liver or aorta, without exclusion of lesions.
3. **Blood-pool-referenced:** log(SUVmax / aortic mean), with blood-pool organs and the heart referenced to the liver.
4. **Marginal normative:** residual of a ridge regression model on age and sex.
5. **Conditional normative, linear:** residual of a ridge regression model on age, sex, and the log median SUV of all other organs on the eroded masks, with the target’s own tissue compartment hidden from the predictors.
6. **Conditional, gradient boosting:** the conditional linear score (method 5) with gradient boosting in place of ridge regression (300 iterations, learning rate 0.05, 15 leaves, at least 20 samples per leaf, L2 penalty 1).
7. **Conditional + aleatoric standard deviation (SD), linear or boosting:** the conditional linear (method 5) or boosting (method 6) score, with the residual divided by a patient-specific SD predicted by a variance model (Eq. 2) instead of an SD that is constant across patients.
8. **Global uptake:** two baselines for the conditional scores and the detection counterparts of M1 (Section 2.3), log SUVmax minus the mean log uptake of the other organs (slope fixed at one) and the residual of a ridge regression model on age, sex, and that mean (learned slope).

**Figure 1.**
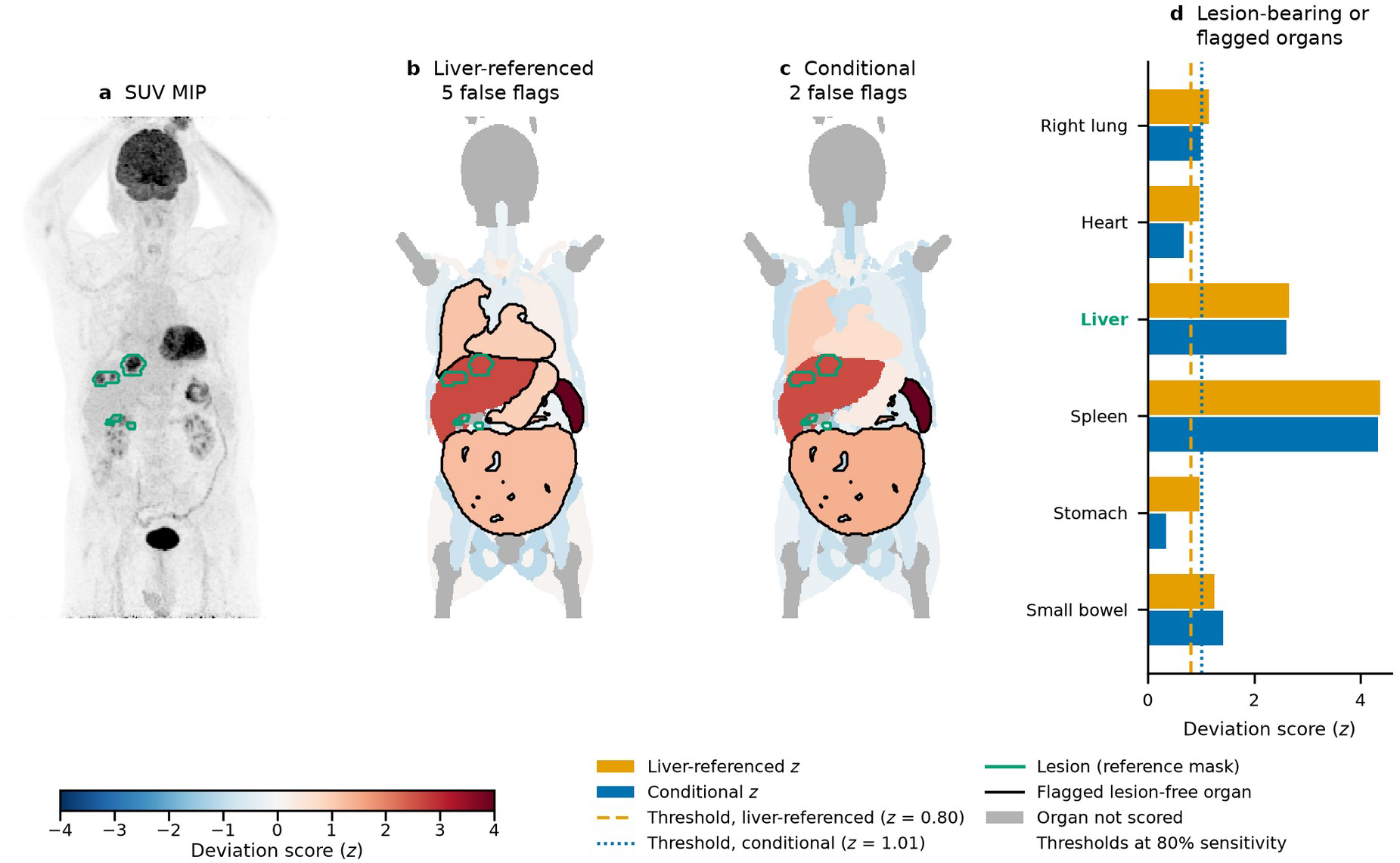
Liver-referenced and conditional deviation scores in one patient. (a) Coronal SUV MIP with the manually segmented lesions released with the dataset (green). (b, c) Organ maps colored by each organ’s deviation score *z*, shown as a coronal projection with soft-tissue organs in front of bone and muscle; gray, organs not scored. Black outlines mark lesion-free organs above each score’s threshold at 80% sensitivity, the operating point of the per-patient analysis (pooled within-patient threshold set on the positives; *z* = 0.80 for liver referencing and 1.01 for the conditional score). (d) *z* of organs that were lesion-bearing (green labels) or above either threshold; dashed and dotted lines, thresholds of the liver-referenced and conditional scores. The case was chosen by a fixed rule: among the 103 patients with one to three lesion-bearing organs, all detected by both scores, it had the median difference in flagged lesion-free organs among those in whom the conditional score flagged fewer. Liver referencing flagged more lesion-free organs than the conditional score in 56% of the 103 patients, as many in 25%, and fewer in 18%. MIP, maximum intensity projection; SUV, standardized uptake value.

All scores were out-of-fold and label-free: no score used the lesion labels, which defined only the positives and negatives of the evaluation. Patients were split into 5 folds, and models were fitted on the control organs of the training folds only. Within the training folds, a second, inner 5-fold split produced out-of-fold residuals for every training control, so that neither the scale of a score nor the variance model was fitted on in-sample residuals. The deviation for organ *o* of patient *i* is

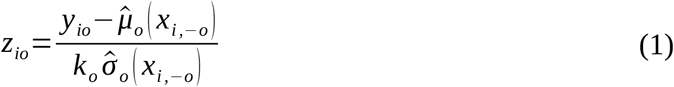

where *y_io_* is the log SUVmax, *x_i_*_,−*o*_ the covariates and other-organ uptake of patient *i* excluding organ *o*, *μ̂_o_* the mean model, *σ̂_o_* a constant (homoscedastic models) or the aleatoric SD, and *k_o_* a factor that rescales the *z* of training controls to unit SD. The aleatoric SD, the patient-specific spread of uptake around the predicted mean [21], came from the variance model, a ridge regression *ĝ_o_* of the log squared inner out-of-fold residual, log *r*^2^, on the same predictors as the mean model, which is the two-step estimator of multiplicative heteroscedasticity [22]:

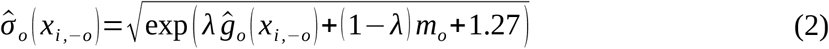

where *λ* = 0.5 shrinks the prediction halfway toward *m_o_*, the mean log squared residual of the training controls, to avoid over-correction, and the constant 1.27 removes the bias of log *r*^2^, because the expectation of the logarithm of a χ^2^_1_ variable is −1.27 [22]. A variant of the boosting score with aleatoric SD added an epistemic term, the spread of a 20-model bootstrap ensemble [23], to the aleatoric variance (Supplementary Table S2).

### 2.5 Evaluation and analysis plan

The primary comparison was the within-patient detection of lesion-bearing organs by the conditional linear score against liver referencing, the clinical standard. A positive was an organ containing at least 2 ml of manually segmented lesion. Negatives were defined in two ways: all organs of controls (against control organs) and the organs without any lesion voxel in patients with malignant lesions (within patients); organs with less than 2 ml of lesion counted as neither. The latter measures specificity in the population in which the score would be used. We report the pooled area under the receiver operating characteristic curve (AUC; Mann–Whitney estimate), the false-positive rate (FPR) at 90% sensitivity, and the per-organ AUC for organs with at least 10 positives and 10 negatives. The FPR was read at 90% sensitivity, the operating point of all pooled comparisons, because an aid that flags organs for review should miss few lesion-bearing organs. Confidence intervals (CIs) for AUC differences came from 1,000 bootstrap resamples of patients. Calibration was the share of control organs with |*z*| > 1.96 (nominal 5%), overall and by within-organ quintile of the predicted aleatoric SD.

Two secondary analyses described the scores at the operating points of a reading aid. First, to express false positives per patient, thresholds for 50–90% sensitivity were set on the positives of the other folds and applied to the held-out fold; we counted the lesion-free organs above threshold in each patient and compared scores by the mean per-patient difference with a patient-level bootstrap CI. Because a reader reviews flags patient by patient, we summarize this burden at 80% sensitivity and also report 70% and 90%. Second, because detection uses only high scores, we report the one-sided share of control organs above z = 1.645, 1.960, 2.326, and 2.576 (nominal 5%, 2.5%, 1%, and 0.5%) and an empirical calibration that converted each *z* to an upper-tail *p* value against the out-of-fold scores of the same organ in the controls of the other folds.

Further secondary analyses tested the robustness of the primary comparison to analysis choices and lesion labels (Section 2.6) and in subgroups with an atypical liver reference (Section 2.10). Others extended it to diagnostic groups (Section 2.7), residual soft-tissue regions and CT covariates (Section 2.8), repeat scans and the number of training controls (Section 2.9), finer organ units (Section 2.11), and individual hotspots, connected components above the PERCIST threshold scored against the expected uptake of the organ or region that contained them (Supplementary Methods and Supplementary Table S8). Confidence intervals of secondary comparisons were not adjusted for multiplicity, and these analyses are interpreted as supporting evidence.

### 2.6 Sensitivity analyses

To test whether the comparison depended on analysis choices, we repeated it with lesion thresholds of 0.5 and 5 ml, without compartment masking, with training on controls plus the lesion-free organs of patients, and with other summaries of the organ’s hottest uptake as the target: SUVpeak, the highest mean SUV in a 1 ml sphere centred in the organ, as defined by PERCIST [6]; the mean of the hottest 1 ml of organ voxels; the mean of the hottest 5%; and the 90th-percentile SUV. Because the maximum of a larger organ is taken over more voxels, we also added the log volume of the target organ to the covariates. The lesion thresholds and compartment masking change which organs count as positive and how much a diffuse compartmental process is explained away, and the extended training set changes the training population. The targets differ in noise and in their sensitivity to small focal lesions: SUVmax is a single voxel, SUVpeak and the hottest 1 ml average the same volume in every organ, and the hottest 5% and the 90th percentile average more voxels in larger organs. Two further analyses tested the dependence on lesion labels: in the first, predictors and reference regions came from the lesion-cleaned masks, which require the manual lesion labels. In the second, a label-free two-pass procedure scored each organ’s eroded median from the other organs and, before the second pass, hid predictor values with *z* > 2.5 (Supplementary Table S6).

### 2.7 Disease groups

To test whether the physiological background of the organs, rather than their lesions, differed by diagnosis, we compared the log lesion-cleaned median SUV of each organ between each diagnostic group and the controls with linear regression adjusted for age and sex, with false discovery rate (FDR) correction over all group-by-organ tests. We also computed, for each group against the controls, the patient-level AUC of two summaries of the deviation scores over a patient’s organs, the maximum and the root mean square of |*z*| (Supplementary Table S3). Finally, we repeated the detection analysis of Section 2.5 separately for each cancer type, with that type’s lesion-bearing organs as positives and, as negatives, the lesion-free organs of patients with the same cancer type (within patients) or all organs of controls (against control organs) (Supplementary Table S5).

### 2.8 Residual soft-tissue regions and CT covariates

Lymph nodes and soft-tissue lesions lie largely outside organ masks. We therefore defined residual soft tissue as the body (CT > −500 Hounsfield units [HU], holes filled, largest component) minus all TotalSegmentator structures (dilated by one voxel), between −200 and 200 HU. Anatomical landmarks split it into six regions: neck (skull base to lung apex), mediastinum (lung apex to liver dome, inside the slice-wise convex hull of the lungs), chest wall and axillae (same levels, outside the hull), abdomen (liver dome to iliac crest), pelvis (iliac crest to femoral heads), and inguinal region (100 mm below the femoral heads). Landmarks were taken from the organ masks after discarding connected components smaller than 10% of the largest and slices with fewer than 20 voxels, because a few mislabeled voxels could otherwise move a landmark by tens of centimeters. When two consecutive landmarks were out of head-to-foot order, the regions on either side were set to missing.

Regions were scored as targets from the organs and covariates, with the median SUV of the whole region as input, because regions are not eroded. Using regions also as predictors was a sensitivity analysis, in which they formed one tissue compartment for masking. A region was retained if it was valid in at least 85% of controls, because the neck is absent when a scan starts below the skull base.

CT covariates were the median HU of the eroded liver, spleen, and paraspinal muscle; whole-body volume and adipose fraction (−190 to −30 HU); and adipose and muscle areas at the level of the third lumbar vertebra (L3). They were winsorized at the 1st and 99th percentiles of the training controls of each fold and added to age and sex as conditioning variables.

### 2.9 Repeat scans and training-set size

Models were trained on the first studies of control patients in five folds over patients, and both the first and the follow-up studies of held-out patients were scored by the same fold model. For organs that were lesion-free at both visits we computed the change in score between visits (Δ*z*), its SD, the intraclass correlation coefficient ICC(C,1), the persistence of flags (*z* > 1.96 at both visits among organs flagged at the first), and the fraction of Δ*z* variance shared by all organs of the same scan pair (the variance of the pair-mean Δ*z* divided by the variance of Δ*z*). The shared fraction measures how much a visit-level shift moves every organ’s score together; with 34 organs, independent organ-level change would give about 0.03. The main analysis used scan pairs without malignant lesions at either visit, and CIs came from patient-level bootstrap resamples. To assess how many controls the models need, each training fold used random subsets of 50, 100, 150, 200, or 250 controls (five subsets each) or all of them, and the same subsets set the mean and SD of the liver-referenced score.

### 2.10 Subgroups with an atypical liver reference

Liver referencing assumes that liver uptake follows the systemic factors that also move other organs. We prespecified three subgroups in which this assumption may fail and compared within-patient detection between liver referencing and the conditional linear model inside each. The first comprised patients with CT signs of hepatic steatosis (liver attenuation below 40 HU or liver minus spleen attenuation below −10 HU); reported effects of steatosis on liver SUV are inconsistent [24, 25], and attenuation criteria are approximate because many autoPET CT scans were contrast-enhanced. The second comprised patients with atypical liver uptake, the top quintile of the absolute conditional *z* of the liver itself, and the third the top tertile of whole-body adipose fraction. For each subgroup we report the AUC difference (ΔAUC) with a patient-level bootstrap CI and the interaction, the difference in ΔAUC between the subgroup and the remaining patients.

### 2.11 Finer organ units

To test whether finer anatomical units add information, we regrouped the TotalSegmentator classes into two finer partitions and repeated the relationship and detection analyses with the same models. The intermediate partition split paired structures into left and right, the lungs into lobes, the venae cavae into superior and inferior, and the sternum from the costal cartilages (56 units, 47 retained by the presence criterion of Section 2.2). The fine partition further separated single vertebrae, ribs by side and level (1–4, 5–8, and 9–12), single gluteal muscles, and single arteries (91 units, 72 retained). Compartment masking was applied as before; every retained unit that came from a subdivided organ belonged to a compartment, so no unit was predicted from another part of its own organ. Relationships were summarized with the predictor sets of Section 2.3 and one additional set, M2s, in which only the other units of the same parent organ were hidden (for example, the other lobes of the same lung) while the rest of the compartment remained available. Comparing M2 with M2s shows how much of a unit’s predictability comes from the remainder of its own organ (Supplementary Table S4). Because finer units change the numbers of positives and negatives, detection was compared at the level of the original organs, on the 29,270 study–organ pairs scored in all partitions, with lesion labels from the original masks. An organ’s score was the maximum *z* over its units, converted to a one-sided *p* value and Šidák-corrected for the number *k* of scored units (1 − (1 − *p*)*^k^*). The uncorrected maximum and an empirical calibration of the maximum against controls of the other folds were secondary aggregations. Within lesion-bearing organs with at least two scored units, we also recorded how often the highest-scoring unit contained lesion.

### 2.12 Reporting and software

Reporting follows the Checklist for Artificial Intelligence in Medical Imaging (CLAIM 2024) [26], and the completed checklist is provided as a separate supplementary file. Analyses used Python 3.11.18 with TotalSegmentator 2.18.0 [19] (nnU-Net v2 2.8.1, PyTorch 2.15.0), scikit-learn 1.9.1 [27], statsmodels 0.15.0, NumPy 2.4.6, pandas 3.1.1, SciPy 1.17.1, nibabel 5.5.0, and Matplotlib 3.11.2; the code is available as described under Code availability.

## 3. Results

### 3.1 Cohort and segmentation quality

Four of the 900 patients lacked age or sex, leaving 896 patients: 412 controls, 167 with lung cancer, 143 with lymphoma, and 174 with melanoma (Table 1). Patients with lymphoma were younger than the other groups (median 47 years). Segmentation succeeded in all 1,014 studies, but the liver-dome index was below 0.7 in 68 studies (6.7%), indicating respiratory PET–CT misregistration at the diaphragm. Adrenals, brain, femurs, and skull had a usable value in fewer than 90% of controls, leaving 34 organs for analysis (Supplementary Table S1); 1.5% of predictor cells were missing and imputed. In the 501 lesion-positive studies, a median 66% of lesion volume lay outside the organ groups.

**Table 1.** Characteristics of the study cohort and lesion-bearing organs by diagnostic group.

| **Group** | **Patients (*n*)** | **Age, median (IQR), y** | **Female, %** | **Organs with ≥ 2 ml of lesion (*n*)** | **Patients with ≥ 2 ml of lesion in at least one organ (*n*)** |
|---|---|---|---|---|---|
| Controls | 412 | 60 (50–70) | 46.6 | – | – |
| Lung cancer | 167 | 65 (59–73) | 38.3 | 854 | 165 |
| Lymphoma | 143 | 47 (29–62) | 47.6 | 547 | 98 |
| Melanoma | 174 | 66 (56–77) | 41.4 | 299 | 92 |
| Total | 896 | 62 (51–72) | 44.2 | 1,700 | 355 |

Each patient is counted once, using the first study with recorded age and sex. Controls are patients without malignant FDG-avid lesions, not healthy volunteers. Lesion-bearing organs are organs containing ≥ 2 ml of manually segmented lesion that were scored by all methods. FDG, 2-deoxy-2-[<sup>18</sup>F]fluoro-D-glucose; IQR, interquartile range.

### 3.2 Inter-organ relationships

Other organs explained most of the between-patient variance in organ uptake. Across 34 organs, the median out-of-fold *R*^2^ was 0.05 with age and sex (M0), 0.34 with global uptake added (M1), and 0.67 with all other organs (M2). Pooled over all organ–patient cells, where the differences between organs made up 76% of the variance, the organ means alone would reach an *R*^2^ of 0.76 and M2 0.89. M2 exceeded M1 for all 34 organs, in all 20 repetitions for 33 of them. Partial correlations formed tissue modules: bone marrow, skeletal muscle, lung, gut, and a visceral module linking liver, blood pool, vessels, and kidneys (Figure 2a). Much of this structure lies within tissue compartments: with the target’s own compartment hidden (M2c), the median *R*^2^ fell to 0.35, close to M1, but stayed above M1 for 25 organs. Cross-compartment information was strong for the liver (0.81), which belongs to no compartment, and for the blood pool (aorta 0.82), and moderate for the kidneys (0.50–0.52) (Figure 2b).

**Figure 2.**
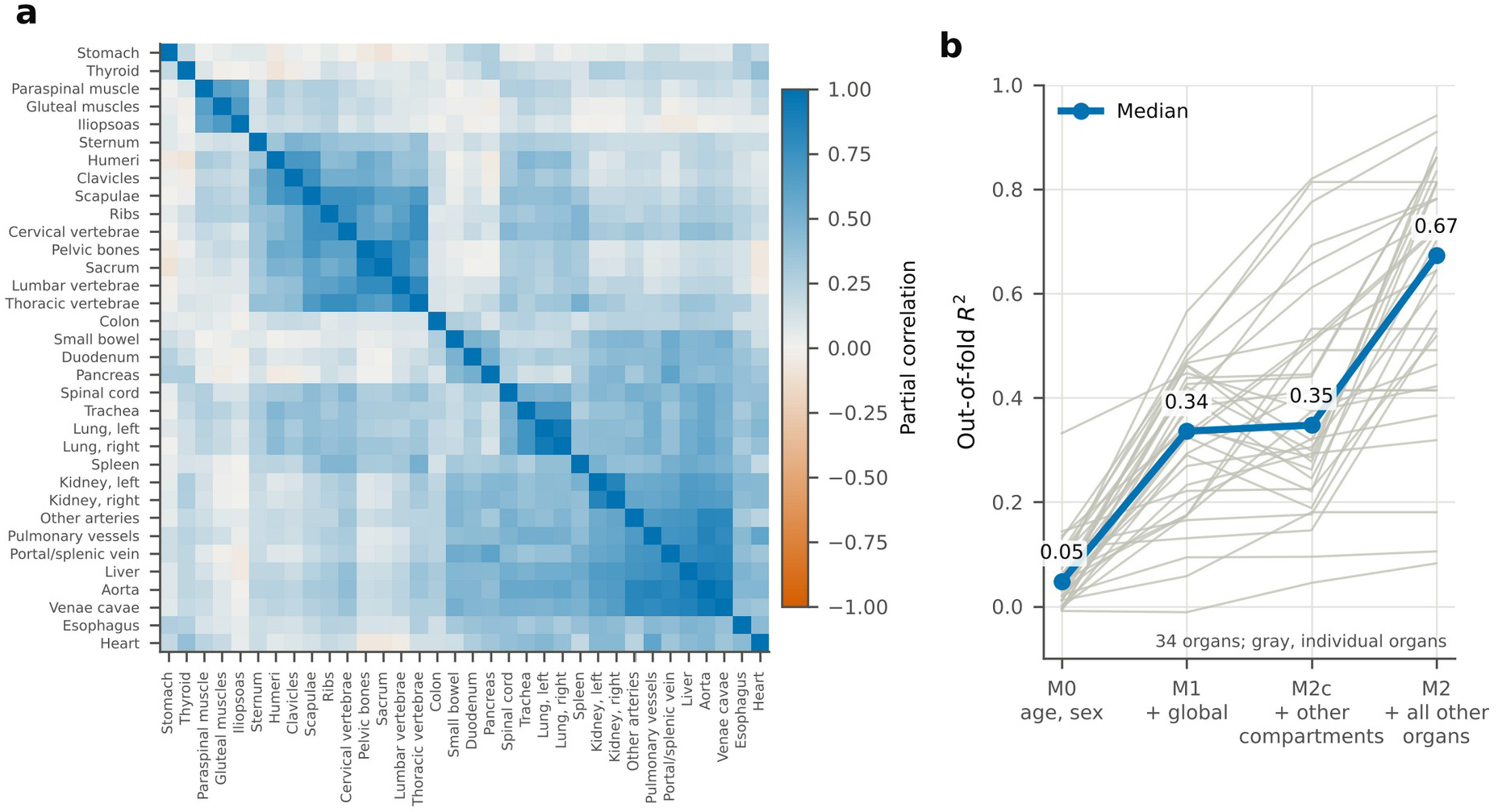
Inter-organ relationships of FDG uptake in control patients (*n* = 412). (a) Partial correlations between organs of the log lesion-cleaned median SUV after removal of age and sex, ordered by hierarchical clustering. (b) Out-of-fold *R*^2^ (5-fold cross-validation repeated 20 times) for predicting each organ’s uptake from four predictor sets: age and sex (M0); M0 plus global uptake (M1); M0 plus organs outside the target’s tissue compartment (M2c); and M0 plus all other organs (M2). Thin gray lines, individual organs (*n* = 34); thick line, median. FDG, 2-deoxy-2-[^18^F]fluoro-D-glucose; SUV, standardized uptake value.

### 3.3 Detection of lesion-bearing organs

The conditional linear model was the best detector (Table 2, Figure 3). Within patients, its AUC was 0.916 against 0.902 for liver referencing (difference 0.014, 95% CI 0.007–0.021), and its FPR at 90% sensitivity was 0.27 against 0.33, a 17% reduction. Against control organs the corresponding values were 0.905 vs 0.894 (difference 0.011, 95% CI 0.004–0.020) and 0.31 vs 0.36. SUVmax, blood-pool referencing, and the marginal normative model did not differ from liver referencing. Neither did a reference on global uptake, the mean of the other organs, whether with a slope fixed at one (AUC 0.902) or learned with age and sex (0.907; ΔAUC 0.005, 95% CI −0.001 to 0.010). The conditional model exceeded the latter by 0.009 (95% CI 0.005 to 0.014), so the gain required organ-specific weights rather than a better global reference.

**Figure 3.**
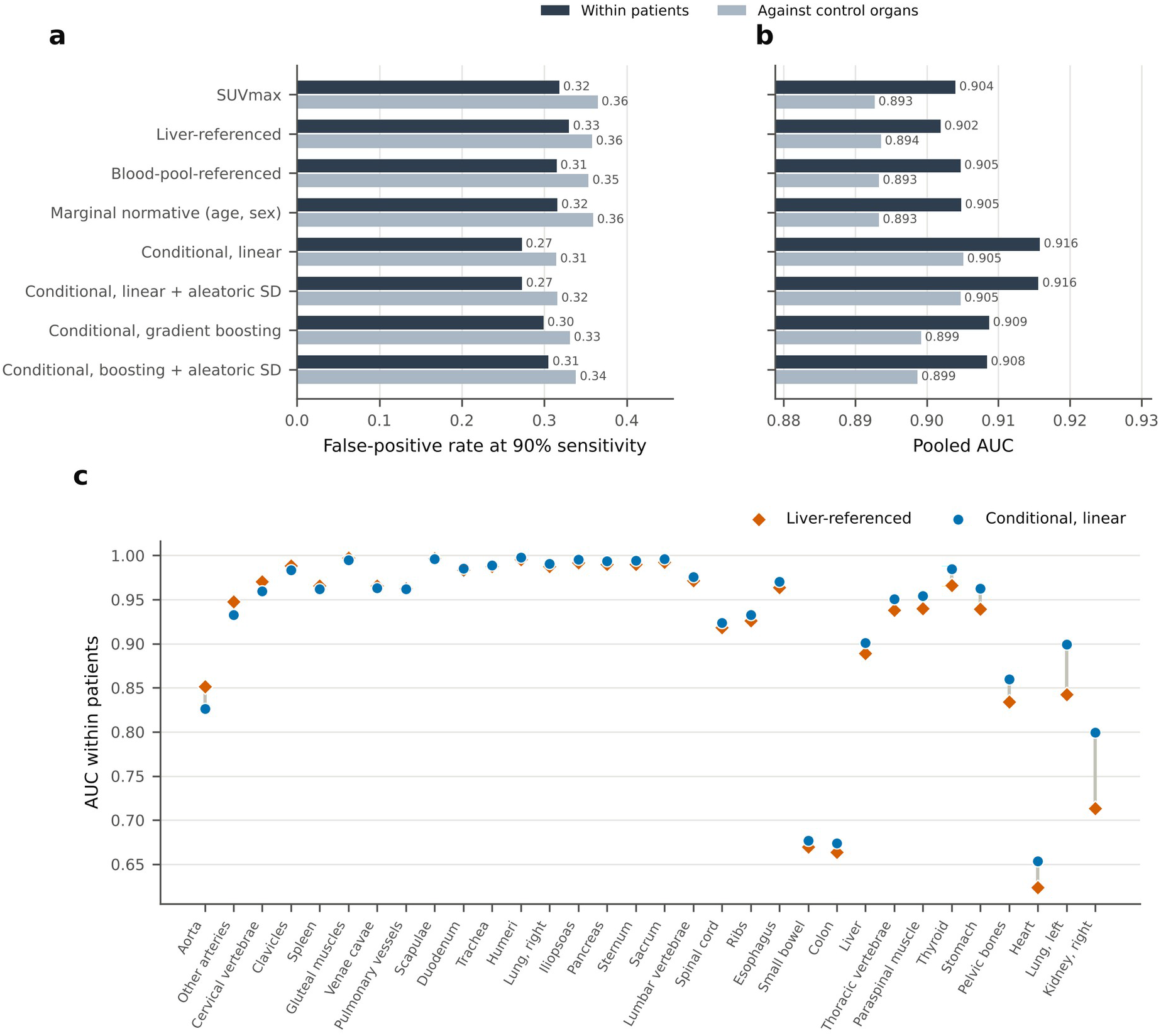
Detection of lesion-bearing organs by each deviation score. (a) False-positive rate at 90% sensitivity and (b) AUC, pooled over organs, for detecting organs containing ≥ 2 ml of lesion. Dark bars use the lesion-free organs of patients as negatives (within patients); light bars use the organs of controls (against control organs). (c) Per-organ AUC within patients for liver-referenced (diamonds) and conditional linear (circles) scores, in organs with at least 10 positives and 10 negatives, sorted by the difference between the two scores. AUC, area under the receiver operating characteristic curve; SD, standard deviation; SUVmax, maximum standardized uptake value.

**Table 2.** Detection of lesion-bearing organs and calibration of deviation scores, pooled over organs.

| Score | Within patients |  |  | Against control organs |  |  | Controls with \|*z*\| > 1.96, % |
|---|---|---|---|---|---|---|---|
|  | AUC | ΔAUC vs liver (95% CI) | FPR at 90% sens. | AUC | ΔAUC vs liver (95% CI) | FPR at 90% sens. |  |
| SUVmax | 0.904 | 0.002 (−0.004 to 0.008) | 0.32 | 0.893 | −0.001 (−0.008 to 0.006) | 0.36 | 4.8 |
| Liver-referenced | 0.902 | – | 0.33 | 0.894 | – | 0.36 | 4.8 |
| Blood-pool-referenced | 0.905 | 0.003 (−0.000 to 0.006) | 0.31 | 0.893 | −0.000 (−0.004 to 0.004) | 0.35 | 4.9 |
| Marginal normative (age, sex) | 0.905 | 0.003 (−0.004 to 0.010) | 0.32 | 0.893 | −0.000 (−0.008 to 0.008) | 0.36 | 4.8 |
| Global uptake, slope fixed at one | 0.902 | 0.000 (−0.005 to 0.005) | 0.32 | 0.892 | −0.002 (−0.007 to 0.004) | 0.35 | 5.0 |
| Global uptake, learned slope (age, sex) | 0.907 | 0.005 (−0.001 to 0.010) | 0.30 | 0.894 | 0.001 (−0.006 to 0.007) | 0.34 | 4.8 |
| **Conditional, linear** | **0.916** | **0.014 (0.007 to 0.021)** | **0.27** | **0.905** | **0.011 (0.004 to 0.020)** | **0.31** | 4.8 |
| Conditional, linear + aleatoric SD | 0.916 | 0.014 (0.007 to 0.021) | 0.27 | 0.905 | 0.011 (0.003 to 0.019) | 0.32 | 5.1 |
| Conditional, gradient boosting | 0.909 | 0.007 (−0.000 to 0.014) | 0.30 | 0.899 | 0.006 (−0.003 to 0.014) | 0.33 | 5.0 |
| Conditional, boosting + aleatoric SD | 0.908 | 0.006 (−0.002 to 0.014) | 0.31 | 0.899 | 0.005 (−0.003 to 0.014) | 0.34 | 5.2 |

Positives are organs containing ≥ 2 ml of lesion (*n* = 1,700). Negatives are the lesion-free organs of patients with malignant lesions (within patients; *n* = 12,547) or all organs of controls (against control organs; *n* = 13,843). ΔAUC is the difference from liver referencing with a patient-level bootstrap 95% CI. Calibration is the share of control organs with \|*z*\| > 1.96 (nominal 5%). Global-uptake scores were computed in a separate run with the same folds; the conditional model exceeded the global-uptake score with learned slope by 0.009 (95% CI 0.005 to 0.014) within patients. Bold, best score. AUC, area under the receiver operating characteristic curve; CI, confidence interval; FPR, false-positive rate; sens., sensitivity; SD, standard deviation; SUVmax, maximum standardized uptake value.

Per patient, the conditional score flagged fewer lesion-free organs at every sensitivity (Figure 4). With thresholds set on the other folds, at 80% sensitivity it flagged 2.90 lesion-free organs per patient against 4.25 with liver referencing (difference −1.35, 95% CI −1.61 to −1.08; fewer in 50% of patients, as many in 33%, and more in 17%), and at 70% sensitivity 1.29 against 2.01 (−0.72, −0.89 to −0.54). At 90% sensitivity every score flagged many organs (7.0 with the conditional score and 8.6 with liver referencing).

**Figure 4.**
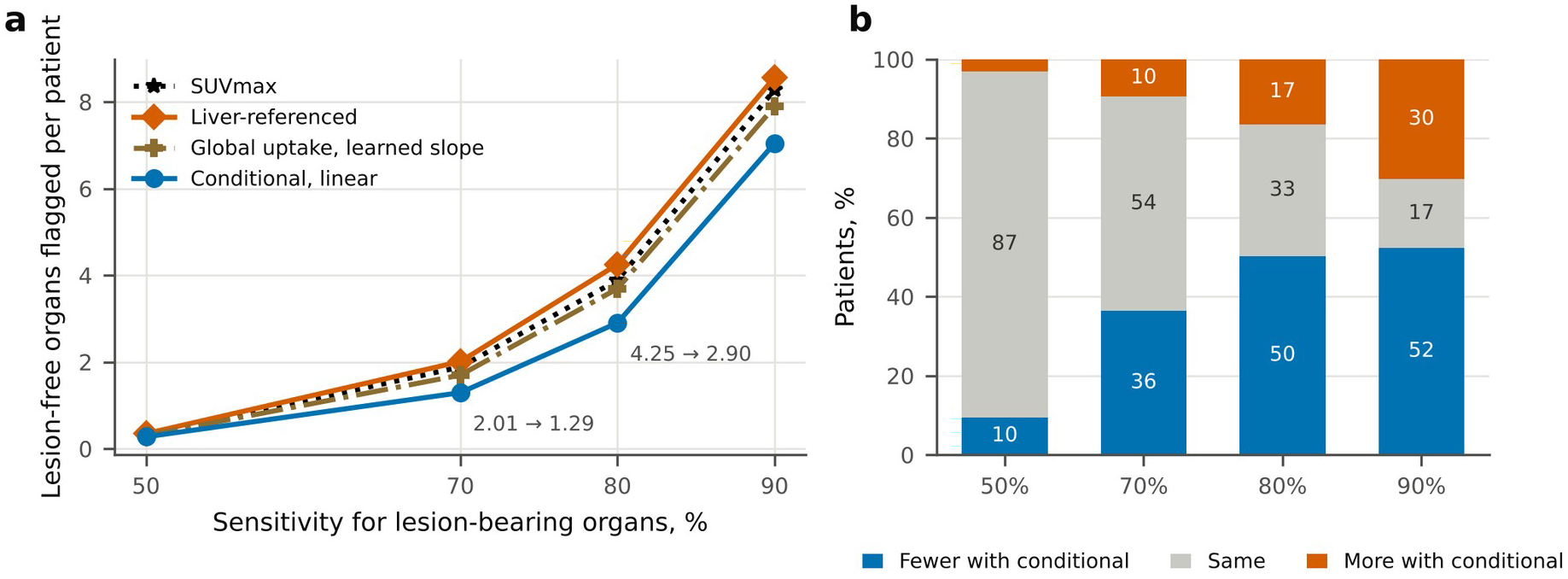
False-positive organs per patient. (a) Mean number of lesion-free organs per patient above the threshold that gives each sensitivity for lesion-bearing organs, with thresholds set on the positives of the other folds; labels give the liver-referenced and conditional values at 70% and 80% sensitivity. Global uptake, learned slope: ridge regression model on age, sex, and the mean uptake of the other organs. (b) Share of patients in whom the conditional linear score flagged fewer, as many, or more lesion-free organs than liver referencing. Within-patient comparison, 484 patients with malignant lesions.

The scores did not depend on lesion labels: eroded and lesion-cleaned medians differed by more than 5% in 3.0% of the patients’ predictor values, and conditional *z* from the two inputs had a rank correlation of 0.998. With lesion-cleaned inputs, the AUCs were 0.904 and 0.918 (ΔAUC 0.013, 95% CI 0.007 to 0.019), and the label-free two-pass procedure gave 0.918 (ΔAUC 0.016, 0.010 to 0.023) (Table 3; Supplementary Table S6). The advantage held in patients with lesions in six or more organs, whose lesions could contaminate the predictors (AUC 0.904 → 0.921).

**Table 3.** Sensitivity of the comparison between the conditional linear model and liver referencing to analysis choices.

| **Setting** | **Positives** | **AUC within patients, liver → conditional** | **ΔAUC (95% CI)** | **FPR at 90% sens., liver → conditional** | **ΔAUC vs control organs (95% CI)** |
|---|---|---|---|---|---|
| Main (≥ 2 ml, SUVmax) | 1,700 | 0.902 → 0.916 | 0.014 (0.007 to 0.021) | 0.33 → 0.27 | 0.011 (0.004 to 0.020) |
| Lesion ≥ 0.5 ml | 2,585 | 0.872 → 0.883 | 0.012 (0.005 to 0.018) | 0.43 → 0.39 | 0.008 (−0.001 to 0.017) |
| Lesion ≥ 5 ml | 1,074 | 0.915 → 0.931 | 0.016 (0.009 to 0.023) | 0.28 → 0.23 | 0.015 (0.006 to 0.023) |
| No compartment masking | 1,700 | 0.902 → 0.915 | 0.013 (0.006 to 0.020) | 0.33 → 0.27 | 0.010 (0.002 to 0.018) |
| Training includes patients' lesion-free organs | 1,700 | 0.902 → 0.913 | 0.011 (0.004 to 0.018) | 0.33 → 0.27 | 0.009 (0.002 to 0.017) |
| Lesion-cleaned inputs (lesion labels used) | 1,700 | 0.904 → 0.918 | 0.013 (0.007 to 0.019) | 0.32 → 0.26 | 0.010 (0.003 to 0.018) |
| Two-pass, label-free | 1,700 | 0.902 → 0.918 | 0.016 (0.010 to 0.023) | 0.33 → 0.26 | 0.014 (0.006 to 0.021) |
| SUVpeak target (1 ml sphere) | 1,700 | 0.898 → 0.916 | 0.018 (0.011 to 0.026) | 0.34 → 0.27 | 0.016 (0.007 to 0.024) |
| Hottest 1 ml target | 1,700 | 0.884 → 0.900 | 0.015 (0.008 to 0.023) | 0.42 → 0.37 | 0.011 (0.002 to 0.020) |
| Hottest 5% target | 1,700 | 0.816 → 0.833 | 0.016 (0.005 to 0.028) | 0.64 → 0.62 | 0.009 (−0.003 to 0.022) |
| 90th-percentile SUV target | 1,700 | 0.695 → 0.697 | 0.002 (−0.016 to 0.022) | 0.81 → 0.82 | −0.010 (−0.032 to 0.011) |
| Organ volume as covariate | 1,700 | 0.902 → 0.916 | 0.014 (0.008 to 0.022) | 0.33 → 0.27 | 0.012 (0.003 to 0.019) |
| CT covariates added | 1,700 | 0.902 → 0.918 | 0.017 (0.010 to 0.024) | 0.33 → 0.27 | 0.014 (0.006 to 0.021) |
| Residual regions as targets | 1,700 | 0.902 → 0.916 | 0.014 (0.007 to 0.021) | 0.33 → 0.27 | 0.011 (0.004 to 0.021) |
| Regions also as predictors | 1,700 | 0.902 → 0.906 | 0.004 (−0.004 to 0.012) | 0.33 → 0.30 | 0.000 (−0.009 to 0.010) |
| Regions also as predictors, mediastinum hidden | 1,700 | 0.902 → 0.917 | 0.015 (0.008 to 0.022) | 0.33 → 0.27 | 0.013 (0.004 to 0.021) |
| Regions as targets and CT covariates | 1,700 | 0.902 → 0.918 | 0.017 (0.009 to 0.024) | 0.33 → 0.27 | 0.014 (0.006 to 0.022) |

Each row changes one setting of the main analysis (lesion ≥ 2 ml, SUVmax target, compartment masking, training on controls only, eroded inputs, no residual regions). Confidence intervals come from separate bootstrap resamples and may differ from Table 2 in the last digit. Region and CT settings report organ-level results; region-level results are in Table 4. Lesion-cleaned inputs use the manual lesion labels to clean predictors and reference regions; the two-pass procedure does not (Section 2.6; Supplementary Table S6). AUC, area under the receiver operating characteristic curve; CI, confidence interval; CT, computed tomography; FPR, false-positive rate; sens., sensitivity; SUV, standardized uptake value; SUVmax, maximum standardized uptake value; SUVpeak, highest mean standardized uptake value in a 1 ml sphere.

### 3.4 Calibration

All deviation scores were calibrated overall, with 4.8–5.2% of control organs at |*z*| > 1.96 (Table 2), but the homoscedastic scores were not uniformly calibrated. In the highest quintile of predicted variability, 7.4% of control organs exceeded the threshold with liver referencing and 6.5% with the conditional linear model, against 3.4–5.0% in the lower quintiles. With the linear model, the variance model flattened this profile (4.9% in the top quintile) at the cost of a slight excess in the lowest quintile (6.8%); with gradient boosting it reduced the excess less (6.5% in the top quintile) (Figure 5).

**Figure 5.**
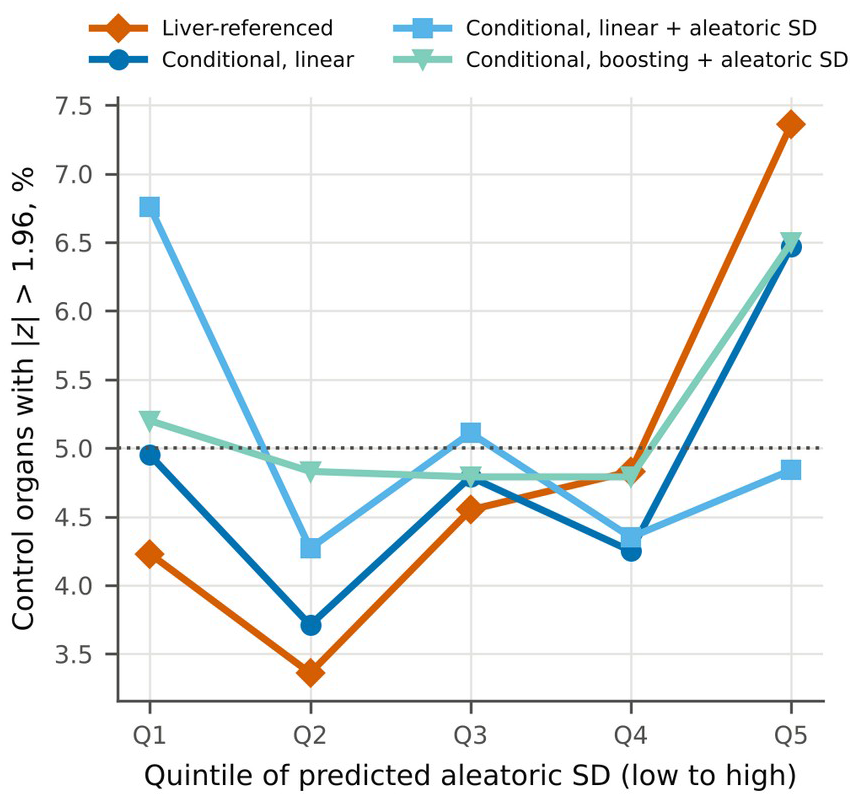
Calibration of deviation scores across levels of predicted physiological variability. Share of control organs with |*z*| > 1.96 in each within-organ quintile of the aleatoric standard deviation (SD) predicted by the variance model (Q1, lowest; Q5, highest). Scores without a variance model (liver-referenced, conditional linear) are grouped by the same quintiles. A calibrated score lies on the dotted line at the nominal 5% in every quintile.

Agreement with the nominal 5% overall hid an asymmetry in the upper tail. Because log SUVmax is a maximum, its deviations were right-skewed (median within-organ skewness of *z* about 1.2), and 6.3% of control organs exceeded *z* = 1.645 with the conditional linear model (nominal 5%), 2.7% exceeded 2.326 (nominal 1%), and 2.0% exceeded 2.576 (nominal 0.5%); liver referencing behaved alike (6.6%, 2.7%, and 2.0%), and only 0.6% and 0.4% of control organs fell below −1.96. Empirical calibration against the controls of the other folds restored the nominal rates (4.9%, 0.9%, and 0.3%) in every organ (4.4–5.5% above *z* = 1.645), at a small cost in pooled ranking (AUC 0.912 vs 0.916), because all *z* values beyond the most extreme control receive the same calibrated value (Supplementary Table S7, Supplementary Figure S1).

### 3.5 Organ-specific gains

Gains concentrated in organs whose physiological uptake varies between patients (Figure 3c). Within patients, the AUC rose from 0.84 to 0.90 for the left lung, from 0.62 to 0.65 for the heart, from 0.71 to 0.80 for the right kidney, from 0.97 to 0.99 for the thyroid, from 0.94 to 0.96 for the stomach, and from 0.83 to 0.86 for the pelvic bones; for the liver, which liver referencing cannot reference to itself, it rose from 0.89 to 0.90. Most bone, muscle, and parenchymal organs had an AUC above 0.95 with both scores. The conditional model was worse for the aorta (0.83 vs 0.85), other arteries (0.93 vs 0.95), and cervical vertebrae (0.96 vs 0.97).

### 3.6 Non-linear models and patient-specific variability

Gradient boosting was slightly worse than the linear model (within patients 0.909 vs 0.916; difference −0.007, 95% CI −0.011 to −0.003). Adding the variance model left the ranking unchanged (0.916 vs 0.916) while making calibration uniform across patients (Section 3.4). Adding the epistemic term from the bootstrap ensemble to the boosting score changed neither detection nor overall calibration (Supplementary Table S2).

### 3.7 Sensitivity analyses

The advantage over liver referencing held for 5 ml lesions, for training that included the lesion-free organs of patients, and without compartment masking (Table 3). It also held with lesion-cleaned inputs and with the two-pass procedure (Section 3.3). At 0.5 ml it remained significant within patients but not against control organs. With SUVpeak as the target, liver referencing lost slightly (AUC 0.898) and the conditional score did not (0.916), so the difference grew to 0.018 (95% CI 0.011 to 0.026) within patients and 0.016 (0.007 to 0.024) against control organs; with the hottest 1 ml it was 0.015 (0.008 to 0.023). Summaries over a share of the organ lowered every score as the share grew (conditional AUC 0.833 for the hottest 5% and 0.697 for the 90th percentile), because in large organs they dilute small lesions. The conditional advantage held for the hottest 5% within patients (0.016, 0.005 to 0.028) but not against control organs (0.009, −0.003 to 0.022), and at the 90th percentile all scores performed poorly and did not differ. Adding organ volume as a covariate left the result unchanged (ΔAUC 0.014, 0.008 to 0.022).

### 3.8 Disease groups

Lesion-cleaned uptake differed between groups on average but did not separate individual patients. Lymphoma showed higher uptake in the spleen (+9.0%) and in vertebral and pelvic marrow (+6.3% to +10.9%) and lower uptake in muscle (−7.5% to −11.0%) than controls, with 16 of 102 tests significant after FDR correction (Supplementary Table S3). Patient-level AUCs for any group against controls were 0.46–0.57 for all scores. By cancer type, the advantage of the conditional score over liver referencing was clear in lung cancer and melanoma but not in lymphoma (Supplementary Table S5). Within patients, the AUC rose from 0.898 to 0.913 in lung cancer (ΔAUC 0.014, 95% CI 0.005 to 0.025) and from 0.922 to 0.946 in melanoma (0.023, 0.009 to 0.042), and the FPR at 90% sensitivity fell from 0.34 to 0.27 and from 0.25 to 0.20. In lymphoma, the AUC changed little (0.921 to 0.926; ΔAUC 0.005, −0.006 to 0.017) and did not differ against control organs (−0.007, −0.019 to 0.004), although the within-patient FPR at 90% sensitivity fell from 0.25 to 0.22.

### 3.9 Residual soft-tissue regions and CT covariates

Landmarks were out of order in 4 of 1,014 studies, whose affected regions were set to missing. The six regions held a median 37% of lesion volume per lesion-positive study, raising coverage together with the organs from 34% to 85%, and contained 926 study-regions with at least 2 ml of lesion. Used as predictors, however, the regions reduced the organ-level gain (within patients 0.902 → 0.906; ΔAUC 0.004, 95% CI −0.004 to 0.012). The mediastinal residual is small (median 294 ml) and contained lesion in 63% of lesion-positive studies, with a median 7.4% of its volume; its median SUV shifted by more than 5% in 36% of lesion-positive studies, against 0.6–1.4% for the other regions and 2.8% for the eroded organ medians. Hiding only the mediastinum from the predictors restored the gain (0.917; ΔAUC 0.015, 0.008 to 0.022), so regions were used as targets only (Table 3).

Pooled over regions, conditional and liver-referenced scores did not differ (within patients 0.831 → 0.838, ΔAUC 0.007, 95% CI −0.002 to 0.015; against control regions ΔAUC 0.008, −0.002 to 0.018), but the pooled figure hid opposite patterns (Table 4). In the mediastinum the conditional score raised the AUC from 0.885 to 0.921 within patients and lowered the FPR at 90% sensitivity from 0.28 to 0.24, and the chest wall and axillae improved slightly. In the neck, abdomen, pelvis, and inguinal region the conditional score was equal or slightly worse. Abdominal, pelvic, and inguinal regions performed poorly for every score (AUC 0.61–0.73). In controls their median SUVmax was 12–20, against 5–7 in the thoracic and neck regions, suggesting that unsegmented physiological uptake (for example bowel wall, ureters, and genital structures) dominates the regional summary.

**Table 4.** Detection of lesion-bearing residual soft-tissue regions with liver-referenced and conditional linear scores.

| **Region** | **Positives** | **AUC vs control regions** | **AUC within patients** | **FPR at 90% sens., within patients** |
|---|---|---|---|---|
| Neck | 128 | 0.901 → 0.894 | 0.923 → 0.913 | 0.23 → 0.25 |
| Mediastinum | 242 | 0.866 → 0.905 | 0.885 → 0.921 | 0.28 → 0.24 |
| Chest wall and axillae | 214 | 0.919 → 0.930 | 0.958 → 0.964 | 0.09 → 0.10 |
| Abdomen | 168 | 0.685 → 0.677 | 0.685 → 0.679 | 0.66 → 0.69 |
| Pelvis | 95 | 0.725 → 0.709 | 0.708 → 0.693 | 0.56 → 0.60 |
| Inguinal region | 79 | 0.679 → 0.671 | 0.609 → 0.612 | 0.73 → 0.72 |
| All regions | 926 | 0.822 → 0.830 | 0.831 → 0.838 | 0.48 → 0.50 |

Positives are regions containing ≥ 2 ml of lesion. Regions were scored as targets from the organs and covariates. Values are given as liver-referenced → conditional linear. Per-region differences were not bootstrapped; only the pooled difference has a CI (Section 3.9). AUC, area under the receiver operating characteristic curve; CI, confidence interval; FPR, false-positive rate; sens., sensitivity.

Adding CT covariates slightly increased the organ-level advantage: within patients the AUC rose from 0.902 to 0.918 (ΔAUC vs liver referencing 0.017, 95% CI 0.010 to 0.024; FPR 0.33 → 0.27), and against control organs ΔAUC was 0.014 (0.006 to 0.021). The gain over the conditional model without CT was small (pooled +0.003) and concentrated in the right kidney, aorta, heart, and cervical vertebrae (+0.010 each) and other arteries (+0.009); the liver changed little (0.901 → 0.903) and muscle did not improve. Calibration was preserved (4.9% of control organs with |*z*| > 1.96; in the highest-SD quintile 7.3% for the linear model and 5.7% with the variance model). With regions as targets and CT covariates combined, the organ-level ΔAUC within patients was 0.017 (0.009 to 0.024).

### 3.10 Repeat scans and training-set size

Eighty-one patients had 114 follow-up studies (median interval 306 days, range 70–2,424), and the main analysis comprised 61 scan pairs in 43 patients without malignant lesions at either visit. About half of the deviation variance persisted for every score (ICC 0.44–0.52; Table 5). The conditional score was slightly less repeatable than liver referencing (SD of Δ*z* 1.03 vs 1.00; difference 0.03, 95% CI 0.00 to 0.06), but its changes were less shared across organs: the shared fraction fell from 0.17 with liver referencing to 0.09 (difference −0.08, 95% CI −0.12 to −0.05), against 0.12 for SUVmax and about 0.03 expected from independent change. A change in liver uptake between visits thus moved every liver-referenced score together, whereas conditional scores changed organ by organ. Of control organs flagged at the first scan, 19% (liver referencing) and 21% (conditional) were flagged again, and results were similar over all 113 pairs.

**Table 5.** Repeat-scan behavior of deviation scores in organs without lesions at either visit.

| **Score** | **ICC(C,1)** | **SD of Δ*z*** | **Shared fraction of Δ*z*** | **Flags persisting** |
|---|---|---|---|---|
| SUVmax | 0.52 | 0.97 | 0.12 | 0.30 |
| Liver-referenced | 0.49 | 1.00 | 0.17 | 0.19 |
| Marginal (age, sex) | 0.51 | 0.97 | 0.12 | 0.29 |
| Conditional, linear | 0.44 | 1.03 | 0.09 | 0.21 |
| Conditional, linear + aleatoric SD | 0.44 | 1.05 | 0.09 | 0.21 |

Analysis of 61 scan pairs from 43 patients without malignant lesions at either visit (2,043 organ pairs; median interval 306 days). All scores have unit SD in controls, so an SD of Δ*z* of 1.41 would mean no relation between visits. Shared fraction, variance of the pair-mean Δ*z* divided by the variance of Δ*z* (about 0.03 if organs change independently); flags persisting, share of organs with *z* > 1.96 at the first scan that exceed 1.96 again at follow-up. Δ*z*, change in deviation score between visits; ICC(C,1), intraclass correlation coefficient (consistency, single measurement); SD, standard deviation; SUVmax, maximum standardized uptake value.

The conditional linear model exceeded liver referencing from 100 training controls onward in every subset (within-patient ΔAUC 0.009 to 0.013) and leveled off at about 200 controls (ΔAUC 0.014; FPR at 90% sensitivity 0.33 → 0.27) (Figure 6). With 50 controls the gain was small and inconsistent (0.001 to 0.010 within patients; −0.002 to 0.008 against control organs). Calibration stayed near nominal at every size (5.6% of control organs with |*z*| > 1.96 with 50 controls and 4.8% with all, about 330 per fold). Gradient boosting started below liver referencing, improved more steeply, passed it at about 200 controls, and remained below the linear model with all controls (within-patient AUC 0.909 vs 0.916), whereas the marginal model did not gain from more controls.

**Figure 6.**
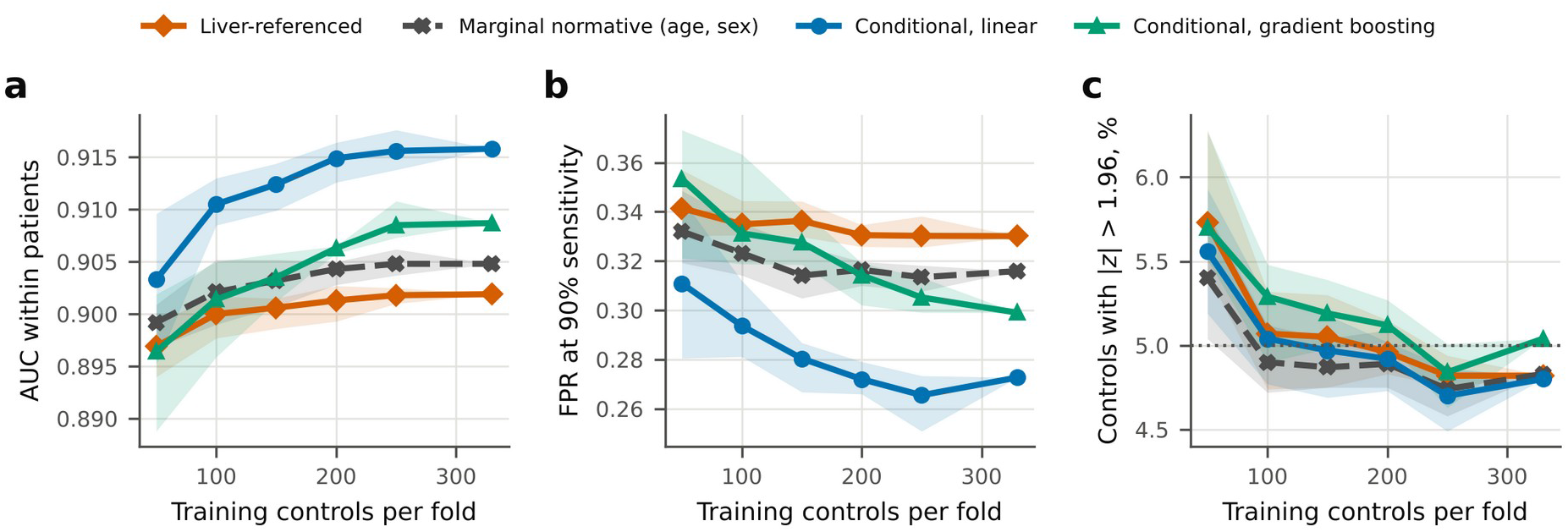
Effect of the number of training controls on detection and calibration. (a) AUC and (b) false-positive rate (FPR) at 90% sensitivity for detecting organs containing ≥ 2 ml of lesion within patients, and (c) share of control organs with |*z*| > 1.96, plotted against the number of training controls per cross-validation fold (50–250, or all, about 330). Lines, mean of five random subsets; shaded bands, range over subsets. The liver-referenced score was standardized on the same subsets. Dotted line in (c), nominal 5%. AUC, area under the receiver operating characteristic curve.

### 3.11 Subgroups with an atypical liver reference

The advantage of the conditional score held in every prespecified subgroup within patients (Table 6). It was numerically larger in patients with CT signs of steatosis (ΔAUC 0.023, 95% CI 0.009 to 0.039, against 0.012 in the other patients), in patients whose liver uptake was atypical for their other organs (0.022, 0.007 to 0.036, against 0.012), and with high adiposity (0.017 against 0.013), but no interaction was significant (all CIs included zero). Against control organs, the gain in the atypical-liver subgroup was not significant (0.015, −0.005 to 0.035).

**Table 6.**
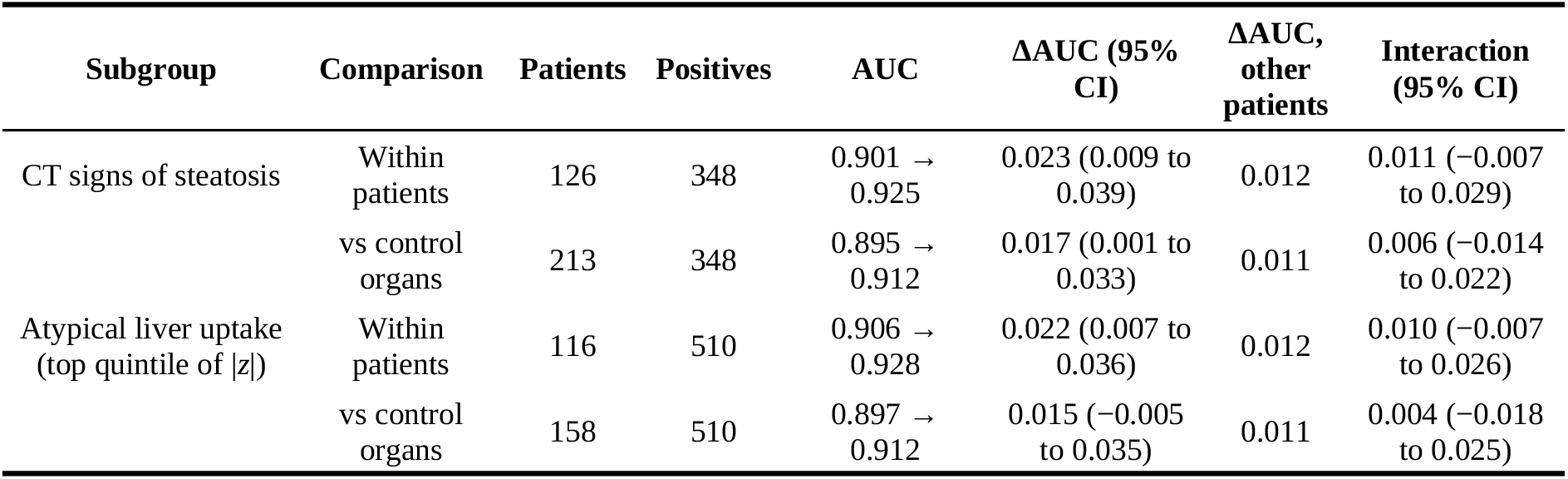
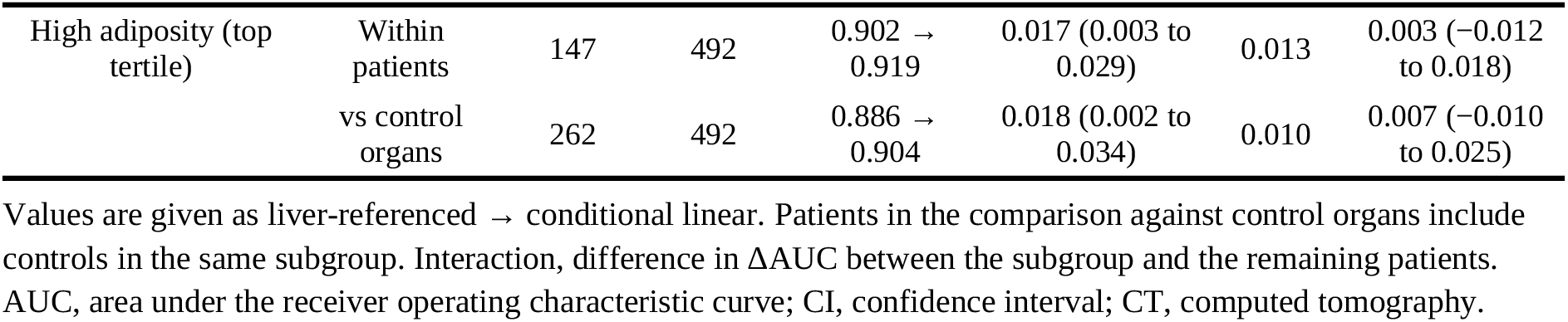
Detection of lesion-bearing organs in prespecified subgroups in which the liver reference may be atypical.

### 3.12 Finer organ units

Finer units raised the out-of-fold *R*^2^ from all other units (median 0.67 with the 34 organs, 0.80 with 47 units, and 0.82 with 72 units), but only because units were predicted from other parts of the same organ: with only those hidden (M2s; Supplementary Table S4), the median *R*^2^ was 0.66 in both finer partitions, and information from outside the unit’s compartment (M2c) fell from 0.35 to 0.30 and 0.25. At the organ level (1,635 lesion-bearing organs; 12,266 lesion-free organs of patients), the Šidák-corrected conditional score performed as with the original organs (AUC 0.915, 0.913, and 0.915; ΔAUC −0.002, 95% CI −0.005 to 0.000, and 0.000, −0.003 to 0.003), and its advantage over liver referencing was retained (ΔAUC 0.015, 0.011, and 0.012; all CIs above zero). Liver referencing and SUVmax gained slightly from finer units (liver referencing, ΔAUC 0.002 and 0.003). The uncorrected maximum lowered every score as units were added (conditional, ΔAUC −0.009 and −0.014), and empirical calibration gave a small loss for the conditional score (−0.005 and −0.004) and none for liver referencing. Within lesion-bearing organs with at least two scored units, the highest-scoring unit contained lesion in 98% (intermediate) and 97% (fine) of organs, against 83% and 75% expected by chance, with similar values for every score.

## 4. Discussion

A normative model that predicts each organ’s uptake from the patient’s other organs detected lesion-bearing organs better than reference-region normalization and reduced the false-positive rate at 90% sensitivity by 17% within patients and the number of falsely flagged organs per patient by about a third at 80% sensitivity. The model is a ridge regression model on summary statistics, trained on 412 control patients, and needs no lesion labels, and the result held across lesion sizes, training sets, and masking schemes.

### 4.1 Reference-region normalization as a special case

Deauville and PERCIST criteria compare uptake with the liver or the blood pool [5, 6], which in regression terms is a conditional normative model with one predictor and a slope fixed at vone. Our conditional model relaxes both restrictions, using every organ and learning the weights. That liver and blood-pool referencing performed no better than raw SUVmax in our data suggests that a single reference carries little of the information that the full set of organs provides. A reference on global uptake, the mean of all other organs, did no better, so the gain comes from weighting each organ for each target rather than from averaging more references. The gains were largest where uptake is driven by patient-specific physiology rather than tumor: myocardium, lung parenchyma, kidney (excretion), thyroid, and stomach. In these organs, other organs and covariates capture part of the physiological state that readers otherwise infer from the image as a whole.

The benefit was not confined to patients in whom the liver itself is a poor reference. It was numerically larger with CT signs of steatosis, atypical liver uptake, or high adiposity, the situations in which liver referencing is most often questioned [24, 25], but without a significant interaction. Across cancer types, the gain was clear in lung cancer and melanoma and small in lymphoma, in which liver referencing already performed well (within-patient AUC 0.92). The gain therefore reflects a general advantage of using many references with learned weights rather than a correction for a few atypical livers.

### 4.2 What the inter-organ structure is

Other organs explained two thirds of the variance in organ uptake, but about half of that came from organs of the same tissue type. Bone marrow sites track each other, as do muscles, while the liver, blood pool, and kidneys form a visceral module, in agreement with group-level descriptions of inter-organ metabolic networks [10–12]. This has a practical consequence: a diffuse process confined to one compartment, such as marrow activation in lymphoma, is partly explained away if the other sites of that compartment are used as predictors. Hiding the target’s compartment, as we did, protects against this without loss of detection.

The residual soft-tissue analysis shows where an organ-level reference stops working. In the mediastinum, where lymph nodes sit in fat with low and stable background uptake, conditioning on the patient’s other organs gave the largest single gain of the study. The same property makes residual regions poor predictors when lesion labels are not used: a few nodes shift the median of the small mediastinal residual, which then distorts the expected uptake of other organs, whereas the median of an eroded organ was hardly affected by lesions. In the abdomen, pelvis, and inguinal region, residual tissue also contains bowel wall, ureters, and genital structures that TotalSegmentator does not label, and a regional SUVmax mostly measures their physiological activity. No choice of reference can rescue such a summary, and finer anatomical units are needed. CT body composition, a plausible proxy for insulin resistance and fasting state, explained little beyond what the other organs already carried, mainly in cardiovascular structures and the kidneys, suggesting that the inter-organ pattern itself captures much of the systemic metabolic state that such covariates are meant to represent.

The repeat scans show what the conditional score removes. About half of an organ’s deviation persisted over a median of ten months for every score, so part of what any deviation score flags in a control is a stable individual trait rather than noise. What differed was the structure of change: with liver referencing, a change in the reference organ between visits, for example from steatosis, medication, or fasting state, shifted all organs together; conditional scores absorbed such shifts and changed organ by organ, at the price of slightly lower repeatability. For follow-up reading, where a change in one organ is the signal of interest, this is the more useful property.

### 4.3 Relation to prior normative and anomaly-detection work

Most normative models in neuroimaging are marginal [7, 8], although self-reconstruction of cortical thickness from other cortical locations showed that conditioning on the same subject improves the link to disease [9]. Our results extend this idea to multi-organ PET and to a direct comparison with the clinical standard. In whole-body PET, individual deviation has so far been expressed in other ways. Network approaches compare a patient’s inter-organ correlation or similarity pattern with a healthy or demographically matched reference [10, 13–15, 28]; they characterize systemic states such as diabetes or pulmonary hypertension, but they score edges or whole patients rather than organs and have not been evaluated for lesion detection. Organ-wise atlases compare each organ’s uptake distribution with that of healthy subjects [29]. A generative model predicts a voxel-wise "normal twin" PET from CT and acquisition attributes and was trained partly on autoPET controls [16]; it is the closest to our aim, but it conditions on anatomy and acquisition rather than on the patient’s uptake elsewhere and was compared with raw and lean-body-mass SUV, not with liver referencing. A whole-body PET/CT foundation model described an atlas of inter-organ correlations in 181 healthy subjects and proposed, but did not test, individual deviation scoring [11].

### 4.4 Organ units and the path to voxel-level deviations

Voxel-level anomaly detection reconstructs a healthy-looking image and flags where the observed image departs from it [30–32]. In whole-body PET these methods localize lesions within organs, which organ-level scores cannot, but they need healthy or image-labeled training data, are sensitive to noise, and do not provide a calibrated, patient-specific range of normal uptake. Their reference is an image that looks normal rather than an expectation derived from the same patient’s physiology, and the organ is a pragmatic unit for such an expectation. Unlike cortical vertices in the brain [9], trunk voxels have no reliable correspondence across patients, whereas automated segmentation provides the same organs in every patient, organ uptake can be estimated robustly despite partial-volume effects and misregistration, and readers and clinical criteria already reason per organ [5, 6]. The cost is spatial detail: a summary such as SUVmax shows that an organ is abnormal but not where, and the failure of the 90th-percentile target shows that small lesions in large organs are invisible to coarse summaries.

The two approaches are therefore complementary rather than competing. Organ-level conditioning answers what a given organ should look like in this patient, and a voxel-level model can answer where within the organ the patient departs from that expectation. A natural next step is a hierarchical model in which the conditional organ estimate, or a learned organ embedding, conditions a decoder that predicts the expected uptake and its variance at each voxel of the same organ, yielding an intra-organ conditional *z*-map calibrated in the same way as the organ scores. Subdividing organs alone does not supply this expectation: with lobes, sides, single vertebrae, and rib levels as units, the added predictive power came from other parts of the same organ, information from other compartments fell, and organ-level detection was unchanged once the maximum over units was corrected for their number, although the highest-scoring unit located the lesion in almost every organ. Finer units thus help to localize abnormal uptake but not to define the expected uptake, which favors conditioning finer spatial models on organ-level estimates rather than replacing them.

Because compartment masking hid the opposite side of paired structures, comparison with the contralateral organ, which readers use, was not tested. Intermediate units such as liver segments and lymph-node stations may still help for the abdominal and pelvic soft tissue in which whole-region summaries failed. The voxel-level lesion masks of autoPET allow such a hierarchical model to be evaluated directly against voxel-level anomaly detection.

### 4.5 Non-linear models and uncertainty

Gradient boosting did not improve on ridge regression: with roughly 400 controls and 34 predictors, the relationships that a flexible model could add are not learnable, and the linear model’s bias is the better trade-off. The learning curve agrees: the linear model leveled off at about 200 controls, whereas gradient boosting was still improving and remained below it with all available controls. A few hundred controls therefore suffice for the linear model, and larger, multi-center control sets or representations learned from images rather than summary statistics are needed before non-linear models can help. The variance model did not change the ranking but corrected a systematic miscalibration: homoscedastic scores, including liver referencing, flag patients with high physiological variability too often. Uniform calibration matters when scores are thresholded per patient, which is how a reader-assist tool would use them. The upper tail needs separate attention: because log SUVmax is a maximum, deviations were right-skewed for every score, and a threshold read from the normal distribution flags two to four times the nominal share of controls beyond *z* ≈ 2.3. Empirical calibration against out-of-fold controls removes this excess and is the better way to set thresholds, whereas the parametric *z* remains the better score for ranking.

### 4.6 Limitations

The main limitation is that all data came from a single center and scanner [17], so the calibration and the gains need external validation; a public test–retest dataset of healthy volunteers acquired on a long-axial-field-of-view scanner [33] and institutional health-screening cohorts offer direct tests of both. The controls were patients without malignant lesions rather than healthy volunteers and may have had inflammation or treatment effects, which makes the specificity estimates conservative but the normative reference imperfect. The repeat scans were not a test–retest design: intervals were long (median ten months) and many follow-up scans came after treatment, so repeatability mixes biological change with measurement noise.

Blood glucose, uptake time, and weight, which affect FDG uptake [3, 4], were not available in the dataset. Such shared factors may produce part of the inter-organ relationships, which are statistical dependencies rather than demonstrated physiological coupling between organs. CT-derived body-composition covariates were a weak substitute, adding about 0.002 in AUC, mostly in cardiovascular structures and the kidneys. The lesion labels covered malignant lesions only, so benign FDG-avid findings in patients counted as negatives, and a lesion touching an organ mask counted as an organ lesion; lymph nodes adjacent to vessels and the heart therefore inflate those organs’ positives. At 90% sensitivity every score still flagged about seven lesion-free organs per patient, and the conditional score about three at 80% sensitivity, so organ-level flags are a screen to prioritize review rather than a stand-alone reading aid. For individual hotspots above the PERCIST threshold, organ-specific normal ranges separated lesions from physiological uptake far better than the liver ratio, but conditioning on other organs added little to them, and with a fixed SUV threshold of 2.5 the unstandardized SUVmax performed best (Supplementary Table S8). Separating hotspots will need models of hotspot features, such as location, size, and CT appearance, rather than of organ-level SUVmax.

Finally, the organ units have limited scope and spatial detail. The 41 groups contained a minority of lesion volume (median 34% per study), and residual soft-tissue regions raised coverage to 85% but extended the conditional advantage only to the mediastinum and chest wall. Organ-level SUVmax does not locate a lesion within an organ, and the masks came from CT-based segmentation, in which respiratory misregistration affected 6.7% of studies; PET-adapted segmentation could reduce such boundary errors [20]. Finer units and voxel-level extensions (Section 4.4) are needed to address these limits.

### 4.7 Implications

Organ-level conditional deviations are cheap to compute from automated segmentation and are calibrated on control patients. They could flag organs for review, provide a patient-specific prior for lesion detection and voxel-level localization, or serve as a label-free pretraining target for whole-body representation learning, in which the same masked-prediction principle is applied to learned organ embeddings.

## 5. Conclusion

Conditioning an organ’s expected FDG uptake on the patient’s other organs generalizes reference-region normalization and reduces false positives in label-free detection of lesion-bearing organs and mediastinal soft tissue. A variance model of patient-specific variability makes these deviation scores uniformly calibrated, and the inter-organ reference can be learned from oncology controls without healthy volunteers. Because organs, unlike trunk voxels, correspond across patients, they are a practical unit for a patient-specific normal reference; extending this reference to voxels within organs and to finer soft-tissue units is the next step toward localizing abnormal uptake.

## Data and Code Availability

### Data availability

This study used only publicly available data. The autoPET whole-body FDG PET/CT dataset (FDG-PET-CT-Lesions; 1,014 studies) and its clinical metadata are openly available as a NIfTI release from the University of Tübingen data repository (https://fdat.uni-tuebingen.de/records/8f14a-pf846) [17]; the original DICOM files are hosted by TCIA (https://doi.org/10.7937/gkr0-xv29) [18].

### Code availability

The image-processing and statistical analysis code is available at https://github.com/hauin/conditional-normative-pet.

## Declarations

### Ethics statement

This study was a retrospective secondary analysis of the publicly available, fully de-identified autoPET dataset (FDG-PET-CT-Lesions; DOI: 10.7937/gkr0-xv29). Collection and public release of the original data were approved by the ethics committee of the contributing institution, as reported in the original data descriptor [17, 18]. The authors used only the publicly released images and metadata, had no access to patient-identifying information, did not contact patients, and did not link the data to any other source. Therefore, no additional institutional review board approval or informed consent was required for this secondary analysis.

### Author contributions

**Chang-hyun Park:** Conceptualization, Methodology, Software, Formal analysis, Investigation, Data curation, Writing original draft, Visualization. **Hae-Jeong Park:** Conceptualization, Methodology, Resources, Writing – review & editing, Supervision, Funding acquisition, Project administration.

**Use of generative AI.** During the preparation of this work, the authors used Claude (Anthropic) as writing and coding assistants; the authors reviewed and verified all content and take full responsibility for it.

## Supporting information

CLAIM 2024 checklist

## Data Availability

This study used only publicly available data. The autoPET whole-body FDG PET/CT dataset (FDG-PET-CT-Lesions; 1,014 studies) and its clinical metadata are openly available as a NIfTI release from the University of Tuebingen data repository (https://fdat.uni-tuebingen.de/records/8f14a-pf846); the original DICOM files are hosted by The Cancer Imaging Archive (TCIA; https://doi.org/10.7937/gkr0-xv29). References: Gatidis S, Hepp T, Frueh M, et al. A whole-body FDG-PET/CT dataset with manually annotated tumor lesions. Sci Data. 2022;9:601. Gatidis S, Kuestner T. A whole-body FDG-PET/CT dataset with manually annotated tumor lesions (FDG-PET-CT-Lesions) [dataset]. The Cancer Imaging Archive; 2022. doi:10.7937/gkr0-xv29.

https://fdat.uni-tuebingen.de/records/8f14a-pf846

https://github.com/hauin/conditional-normative-pet

## Acknowledgments

The authors thank the autoPET organizers, the University Hospital Tübingen, and The Cancer Imaging Archive for making the data available.

