## Supplementary Material (Supplementary Methods, Tables S1-S8, Figure S1) for "Conditional normative modeling of whole-body FDG PET/CT: inter-organ relationships outperform reference-region normalization for label-free detection of lesion-bearing organs"

#### Contents


- **Supplementary Methods**: lesion-level (hotspot) analysis
- **Supplementary Table S1**: organ groups and tissue compartments
- **Supplementary Table S2**: detection and calibration of the conditional scores with aleatoric and epistemic SD
- **Supplementary Table S3**: disease groups
- **Supplementary Table S4**: finer organ units
- **Supplementary Table S5**: detection by cancer type
- **Supplementary Table S6**: dependence on lesion labels
- **Supplementary Table S7**: upper-tail calibration
- **Supplementary Table S8**: lesion-level (hotspot) analysis
- **Supplementary Figure S1**: distribution of deviation scores in control organs


All scores use eroded-mask inputs without lesion labels unless stated otherwise.

The completed CLAIM 2024 checklist is provided as a separate file.

### Supplementary Methods: lesion-level (hotspot) analysis

Candidate hotspots were found without lesion labels in the first study of each patient. The liver reference was the mean and SD of SUV in the eroded liver mask, and voxels above the PERCIST threshold (1.5 × liver mean + 2 × liver SD) formed candidates; a fixed threshold of SUV 2.5 was a secondary definition. Voxels in the brain and urinary bladder were excluded, components were defined with 26-connectivity, and components smaller than 0.5 ml were discarded. Each hotspot was assigned to the organ group holding at least half of its voxels or, for hotspots mostly outside the organ groups, to the residual soft-tissue region holding at least half of them; other hotspots and those in organs not analyzed were left unscored.

The manual lesion masks were used only to label hotspots: a hotspot was a lesion if at least 50% of its voxels were lesion and non-lesion if none were; the others were excluded. For each held-out patient and host, the expected log SUVmax and its scale came from the same out-of-fold models as the organ-level scores, and a hotspot's score was (log SUVmax of the hotspot − expected log SUVmax of the host) / scale. Unstandardized scores were the hotspot SUVmax and the ratio of hotspot SUVmax to the liver mean, as in PERCIST. Lesion and non-lesion hotspots of the same patients were compared with the AUC, and differences between scores had patient-level bootstrap 95% CIs (1,000 resamples).

**Supplementary Table S1.** Organ groups merged from TotalSegmentator v2 classes, their tissue compartments for compartment masking, and their inclusion in the analysis.

| Organ group | TotalSegmentator classes | Compartment | Status |
|---|---|---|---|
| Brain | brain | None | Excluded (usable in < 90% of controls) |
| Thyroid | thyroid_gland | None | Analyzed |
| Spinal cord | spinal_cord | None | Analyzed |
| Left lung | lung_upper_lobe_left, lung_lower_lobe_left | Lung and airway | Analyzed |
| Right lung | lung_upper_lobe_right, lung_middle_lobe_right, lung_lower_lobe_right | Lung and airway | Analyzed |
| Trachea | trachea | Lung and airway | Analyzed |
| Esophagus | esophagus | Gastrointestinal tract | Analyzed |
| Heart | heart | None | Analyzed |
| Aorta | aorta | Blood pool and vessels | Analyzed |
| Pulmonary vessels | pulmonary_artery, pulmonary_vein | Blood pool and vessels | Analyzed |
| Venae cavae | superior_vena_cava, inferior_vena_cava | Blood pool and vessels | Analyzed |
| Other arteries | brachiocephalic_trunk, subclavian_artery_left/right, common_carotid_artery_left/right, iliac_artery_left/right | Blood pool and vessels | Analyzed |
| Portal and splenic vein | portal_vein_and_splenic_vein | Blood pool and vessels | Analyzed |
| Liver | liver | None | Analyzed |
| Spleen | spleen | None | Analyzed |
| Pancreas | pancreas | None | Analyzed |
| Stomach | stomach | Gastrointestinal tract | Analyzed |
| Duodenum | duodenum | Gastrointestinal tract | Analyzed |
| Small bowel | small_bowel | Gastrointestinal tract | Analyzed |
| Colon | colon | Gastrointestinal tract | Analyzed |
| Gallbladder | gallbladder | None | Excluded a priori (often removed) |
| Adrenal glands | adrenal_gland_left/right | None | Excluded (usable in < 90% of controls) |
| Left kidney | kidney_left | Kidney | Analyzed |
| Right kidney | kidney_right | Kidney | Analyzed |
| Urinary bladder | urinary_bladder | None | Excluded a priori (excretion) |
| Prostate | prostate | None | Excluded a priori (sex-specific) |
| Skull | skull | Bone | Excluded (usable in < 90% of controls) |
| Cervical vertebrae | vertebrae_C1–C7 | Bone | Analyzed |
| Thoracic vertebrae | vertebrae_T1–T12 | Bone | Analyzed |
| Lumbar vertebrae | vertebrae_L1–L5 | Bone | Analyzed |
| Sacrum | sacrum, vertebrae_S1 | Bone | Analyzed |
| Ribs | rib_left/right_1–12 | Bone | Analyzed |
| Sternum | sternum, costal_cartilages | Bone | Analyzed |
| Scapulae | scapula_left/right | Bone | Analyzed |
| Clavicles | clavicula_left/right | Bone | Analyzed |
| Humeri | humerus_left/right | Bone | Analyzed |

| | | | |
|---|---|---|---|
| Hip bones | hip_left/right | Bone | Analyzed |
| Femurs | femur_left/right | Bone | Excluded (usable in < 90% of controls) |
| Autochthonous back muscles | autochthon_left/right | Skeletal muscle | Analyzed |
| Iliopsoas muscles | iliopsoas_left/right | Skeletal muscle | Analyzed |
| Gluteal muscles | gluteus_maximus/medius/minimus_left/right | Skeletal muscle | Analyzed |

Compartment masking hid all organs of the target's compartment from the predictors of the conditional models; organs without a compartment (None) were predicted from all other organs. Organs were excluded a priori or when a usable eroded-mask value (not truncated by the scan range, at least 20 voxels) was available in fewer than 90% of controls, leaving 34 organs. The two kidneys were kept as separate groups; paired structures otherwise formed one group.

**Supplementary Table S2.** Detection of lesion-bearing organs and calibration of conditional deviation scores with and without patient-specific (aleatoric) and model (epistemic) uncertainty.

| **Score** | **AUC, within patients** | **FPR at 90% sens., within patients** | **AUC, vs control organs** | **FPR at 90% sens., vs control organs** | **Controls with \|*z*\| > 1.96, %** | **Q1, %** | **Q2, %** | **Q3, %** | **Q4, %** | **Q5, %** |
|---|---|---|---|---|---|---|---|---|---|---|
| Conditional, linear | 0.916 | 0.27 | 0.905 | 0.31 | 4.8 | 5.0 | 3.7 | 4.8 | 4.2 | 6.5 |
| Conditional, linear + aleatoric SD | 0.916 | 0.27 | 0.905 | 0.32 | 5.1 | 6.8 | 4.3 | 5.1 | 4.3 | 4.8 |
| Conditional, boosting | 0.909 | 0.30 | 0.899 | 0.33 | 5.0 | 4.2 | 4.0 | 4.4 | 5.1 | 7.7 |
| Conditional, boosting + aleatoric SD | 0.908 | 0.31 | 0.899 | 0.34 | 5.2 | 5.2 | 4.8 | 4.8 | 4.8 | 6.5 |
| Conditional, boosting + aleatoric and epistemic SD | 0.908 | 0.31 | 0.898 | 0.34 | 5.0 | 5.0 | 4.9 | 4.7 | 4.8 | 5.9 |

The aleatoric SD came from the variance model (Eq. 2 of the main text); the epistemic term added the variance of a 20-model bootstrap ensemble of the boosting model to the aleatoric variance. Q1–Q5, share of control organs with $|z| > 1.96$ in each within-organ quintile of the predicted aleatoric SD (Q1, lowest; nominal 5%). Positives are organs with ≥ 2 ml of lesion ($n$ = 1,700); negatives as in Table 2 of the main text. AUC, area under the receiver operating characteristic curve; FPR, false-positive rate; SD, standard deviation; sens., sensitivity.

**Supplementary Table S3.** Disease groups. (a) Organs whose lesion-cleaned uptake differed from that of controls after false discovery rate (FDR) correction. (b) Patient-level separation of each diagnostic group from controls by summaries of the deviation scores.

**(a)**

| Group | Organ | Difference from controls, % | FDR-adjusted *q* |
|---|---|---|---|
| Lung cancer | Left lung | −10.0 | 0.003 |
| Lung cancer | Esophagus | −8.4 | 0.004 |
| Lung cancer | Heart | −5.9 | 0.037 |
| Lymphoma | Gluteal muscles | −11.0 | < 0.001 |
| Lymphoma | Autochthonous back muscles | −10.8 | < 0.001 |
| Lymphoma | Thyroid | −9.1 | 0.017 |
| Lymphoma | Iliopsoas muscles | −7.5 | 0.008 |
| Lymphoma | Heart | −7.5 | 0.012 |
| Lymphoma | Esophagus | −6.7 | 0.049 |
| Lymphoma | Cervical vertebrae | +6.3 | 0.040 |
| Lymphoma | Thoracic vertebrae | +7.5 | 0.019 |
| Lymphoma | Spleen | +9.0 | 0.002 |
| Lymphoma | Lumbar vertebrae | +10.0 | 0.004 |
| Lymphoma | Hip bones | +10.5 | 0.006 |
| Lymphoma | Sacrum | +10.9 | 0.013 |
| Melanoma | Gluteal muscles | −5.7 | 0.013 |

Linear regression of the log lesion-cleaned median SUV on group, age, and sex in each organ (102 tests: 34 organs × 3 groups); the difference is the back-transformed group coefficient. Only tests with $q < 0.05$ are listed.

**(b)**

| Group | Summary over organs | Patients | AUC, marginal | AUC, conditional | ΔAUC (95% CI) |
|---|---|---|---|---|---|
| Lung cancer | Maximum $\|z\|$ | 167 | 0.465 | 0.525 | 0.060 (0.021 to 0.103) |
| Lung cancer | Root mean square of $\|z\|$ | 167 | 0.479 | 0.521 | 0.042 (−0.004 to 0.085) |
| Lymphoma | Maximum $\|z\|$ | 143 | 0.549 | 0.550 | 0.001 (−0.034 to 0.039) |
| Lymphoma | Root mean square of $\|z\|$ | 143 | 0.564 | 0.570 | 0.006 (−0.037 to 0.048) |
| Melanoma | Maximum $\|z\|$ | 174 | 0.561 | 0.540 | −0.021 (−0.059 to 0.014) |
| Melanoma | Root mean square of $\|z\|$ | 174 | 0.547 | 0.524 | −0.024 (−0.070 to 0.016) |

Each group was compared with the 412 controls. Marginal, age–sex normative score; conditional, conditional linear score. ΔAUC, conditional minus marginal, with a patient-level bootstrap 95% CI. An AUC near 0.5 means that the summary did not separate the group from controls. AUC, area under the receiver operating characteristic curve; CI, confidence interval.

**Supplementary Table S4.** Inter-organ relationships and organ-level detection with finer anatomical units.

| Partition | Units, defined / retained | Median $R^2$, M2 | Median $R^2$, M2s | Median $R^2$, M2c | AUC, liver-referenced | AUC, conditional | ΔAUC conditional vs original organs (95% CI) | ΔAUC conditional vs liver (95% CI) | AUC, conditional, uncorrected maximum | Lesion in top unit, % (chance) |
|---|---|---|---|---|---|---|---|---|---|---|
| Original organs | 41 / 34 | 0.67 | – | 0.35 | 0.900 | 0.915 | – | 0.015 (0.008 to 0.022) | 0.915 | – |
| Intermediate | 56 / 47 | 0.80 | 0.66 | 0.30 | 0.902 | 0.913 | −0.002 (−0.005 to 0.000) | 0.011 (0.004 to 0.018) | 0.906 | 98 (83) |
| Fine | 91 / 72 | 0.82 | 0.66 | 0.25 | 0.903 | 0.915 | 0.000 (−0.003 to 0.003) | 0.012 (0.006 to 0.019) | 0.901 | 97 (75) |

M2, all other units; M2s, all other units except those of the same parent organ (for example, the other lobes of the same lung); M2c, units outside the target's tissue compartment. Detection was evaluated at the level of the original organs within patients, on 29,270 study–organ pairs scored in all partitions (1,635 lesion-bearing organs; 12,266 lesion-free organs of patients). An organ's score was the maximum $z$ over its units, Šidák-corrected for the number of scored units, except in the uncorrected-maximum column. Lesion in top unit, share of lesion-bearing organs with at least two scored units in which the highest-scoring conditional unit contained lesion, with the share expected by chance in parentheses. AUC, area under the receiver operating characteristic curve; CI, confidence interval.

**Supplementary Table S5.** Detection of lesion-bearing organs by cancer type.

| Cancer type | Positives (patients) | Score | AUC, within patients | FPR at 90% sens., within patients | ΔAUC vs liver, within patients (95% CI) | AUC, vs control organs | FPR at 90% sens., vs control organs | ΔAUC vs liver, vs control organs (95% CI) |
|---|---|---|---|---|---|---|---|---|
| Lung cancer | 854 (165) | SUVmax | 0.899 | 0.33 | | 0.860 | 0.46 | |
| | | Liver-referenced | 0.898 | 0.34 | | 0.861 | 0.46 | |
| | | Blood-pool-referenced | 0.903 | 0.32 | | 0.857 | 0.47 | |
| | | Marginal normative (age, sex) | 0.904 | 0.32 | | 0.866 | 0.45 | |
| | | Conditional, linear | 0.913 | 0.27 | 0.015 (0.005 to 0.025) | 0.879 | 0.41 | 0.018 (0.006 to 0.030) |
| | | Conditional, linear + aleatoric SD | 0.913 | 0.27 | | 0.880 | 0.41 | |
| Lymphoma | 547 (98) | SUVmax | 0.920 | 0.26 | | 0.923 | 0.26 | |
| | | Liver-referenced | 0.921 | 0.25 | | 0.932 | 0.21 | |
| | | Blood-pool-referenced | 0.923 | 0.25 | | 0.932 | 0.21 | |
| | | Marginal normative (age, sex) | 0.913 | 0.31 | | 0.913 | 0.32 | |
| | | Conditional, linear | 0.926 | 0.22 | 0.005 (−0.006 to 0.017) | 0.925 | 0.23 | −0.007 (−0.019 to 0.004) |
| | | Conditional, linear + aleatoric SD | 0.925 | 0.22 | | 0.923 | 0.24 | |
| Melanoma | 299 (92) | SUVmax | 0.935 | 0.20 | | 0.932 | 0.21 | |
| | | Liver-referenced | 0.922 | 0.25 | | 0.916 | 0.25 | |
| | | Blood-pool-referenced | 0.934 | 0.23 | | 0.927 | 0.24 | |
| | | Marginal normative (age, sex) | 0.939 | 0.19 | | 0.935 | 0.19 | |
| | | Conditional, linear | 0.946 | 0.20 | 0.023 (0.009 to 0.042) | 0.943 | 0.20 | 0.027 (0.012 to 0.046) |
| | | Conditional, linear + aleatoric SD | 0.946 | 0.19 | | 0.943 | 0.19 | |

Positives are the organs with ≥ 2 ml of lesion of patients with the given cancer type (number of patients in parentheses). Within patients, negatives are the lesion-free organs of patients with the same cancer type; against control organs, negatives are all organs of the 412 controls. ΔAUC, conditional linear minus liver-referenced AUC with a patient-level bootstrap 95% CI (1,000 resamples). Scores are the out-of-fold scores of the main analysis (eroded inputs). AUC, area under the receiver operating characteristic curve; CI, confidence interval; FPR, false-positive rate; SD, standard deviation; sens., sensitivity; SUVmax, maximum standardized uptake value.

**Supplementary Table S6.** Dependence of the comparison on lesion labels. (a) Inputs from eroded masks without lesion labels (main analysis), from lesion-cleaned masks, which require the lesion labels, and from a label-free two-pass procedure. (b) Detection by the number of organs with any lesion in the patient (eroded inputs).

**(a)**

| Inputs | AUC within patients, liver → conditional | ΔAUC within patients (95% CI) | FPR at 90% sens., liver → conditional | ΔAUC vs control organs (95% CI) | AUC, global uptake (ridge) | Conditional minus global uptake, within patients (95% CI) |
|---|---|---|---|---|---|---|
| Eroded masks, no lesion labels (main) | 0.902 → 0.916 | 0.014 (0.007 to 0.021) | 0.33 → 0.27 | 0.011 (0.004 to 0.019) | 0.907 | 0.009 (0.004 to 0.014) |
| Lesion-cleaned masks (lesion labels used) | 0.904 → 0.918 | 0.013 (0.007 to 0.019) | 0.32 → 0.26 | 0.010 (0.003 to 0.018) | 0.907 | 0.010 (0.006 to 0.015) |
| Two-pass, label-free | 0.902 → 0.918 | 0.016 (0.010 to 0.023) | 0.33 → 0.26 | 0.014 (0.006 to 0.021) | 0.907 | 0.011 (0.006 to 0.015) |

Lesion-cleaned inputs use the manual lesion masks to remove lesions (dilated by 4 mm) from the predictors and reference regions; the scores are otherwise identical. The two-pass procedure first scored each organ's eroded median from the other organs (conditional linear, out of fold) and set predictor values with $z > 2.5$ to missing before the second pass; it hid 1.0% of control and 1.5% of patient predictor values, including 3.6% of values from organs that contained lesion. Eroded and lesion-cleaned medians differed by more than 5% in 3.0% and by more than 20% in 0.7% of the patients' predictor values. Global uptake (ridge), a ridge regression model on age, sex, and the mean log uptake of the other organs.

**(b)**

| Organs with lesion in the patient | Patients | Positives | Negatives | AUC within patients, liver → conditional | FPR at the pooled 90%-sensitivity threshold, liver → conditional |
|---|---|---|---|---|---|
| 0 | 63 | 0 | 2,129 | – | 0.35 → 0.31 |
| 1–2 | 66 | 35 | 2,131 | 0.880 → 0.920 | 0.33 → 0.29 |
| 3–5 | 92 | 108 | 2,742 | 0.861 → 0.893 | 0.32 → 0.27 |
| ≥ 6 | 263 | 1,557 | 5,545 | 0.904 → 0.921 | 0.33 → 0.25 |

Patients with malignant lesions grouped by the number of organs containing any lesion voxel, which is the number of predictors a lesion could contaminate. Patients without any organ with ≥ 2 ml of lesion have no positives; for them only the false-positive rate at the threshold of the pooled analysis is given. AUC, area under the receiver operating characteristic curve; CI, confidence interval; FPR, false-positive rate; sens., sensitivity.

**Supplementary Table S7.** Upper-tail calibration of deviation scores in control organs.

| **Score** | **\|*z*\| > 1.96, % (5)** | ***z* > 1.645, % (5)** | ***z* > 1.960, % (2.5)** | ***z* > 2.326, % (1)** | ***z* > 2.576, % (0.5)** | ***z* < −1.96, % (2.5)** | **Skewness, median over organs** | **Range over organs, *z* > 1.645, %** | **AUC within patients** |
|---|---|---|---|---|---|---|---|---|---|
| SUVmax | 4.8 | 6.1 | 4.0 | 2.5 | 1.8 | 0.8 | 0.93 | 3.9–8.6 | 0.904 |
| Liver-referenced | 4.8 | 6.6 | 4.4 | 2.7 | 2.0 | 0.4 | 1.21 | 4.1–9.2 | 0.902 |
| Blood-pool-referenced | 4.9 | 6.4 | 4.5 | 2.6 | 1.9 | 0.4 | 1.28 | 3.7–9.2 | 0.905 |
| Marginal normative (age, sex) | 4.8 | 6.0 | 4.0 | 2.5 | 1.8 | 0.8 | 0.98 | 3.6–8.2 | 0.905 |
| Global uptake, slope fixed at one | 5.0 | 6.3 | 4.3 | 2.7 | 1.9 | 0.7 | 1.26 | 3.4–8.2 | 0.902 |
| Global uptake, learned slope (age, sex) | 4.8 | 6.3 | 4.2 | 2.7 | 2.0 | 0.6 | 1.26 | 4.1–8.0 | 0.907 |
| Conditional, linear | 4.8 | 6.3 | 4.2 | 2.7 | 2.0 | 0.6 | 1.24 | 4.4–8.2 | 0.916 |
| Conditional, linear + aleatoric SD | 5.1 | 6.7 | 4.5 | 3.0 | 2.3 | 0.6 | 1.19 | 4.4–9.2 | 0.916 |
| Liver-referenced, empirically calibrated | 4.8 | 4.9 | 2.4 | 1.0 | 0.3 | 2.4 | 0.00 | 4.2–5.4 | 0.895 |
| Conditional, linear, empirically calibrated | 4.9 | 4.9 | 2.4 | 0.9 | 0.3 | 2.5 | 0.00 | 4.4–5.5 | 0.912 |
| Conditional, linear + aleatoric SD, empirically calibrated | 4.9 | 4.9 | 2.5 | 1.0 | 0.3 | 2.5 | 0.00 | 4.2–5.8 | 0.912 |

Nominal rates in parentheses. Shares are pooled over 13,843 control organs (412 controls). Empirical calibration converted each $z$ to an upper-tail $p$ value against the out-of-fold scores of the same organ in the controls of the other folds, $p = (1 + \text{number of control scores} \geq z) / (\text{number of control scores} + 2)$, and back to a standard-normal quantile. Its lower tail is symmetric by construction, and scores beyond the largest control score are tied, which lowers the pooled AUC. The global-uptake and empirically calibrated scores come from a separate run with the same folds. AUC, area under the receiver operating characteristic curve.

**Supplementary Table S8.** Lesion-level analysis: separation of lesion hotspots from non-lesion hotspots in the same patients.

| Candidates | Hosts | Hotspots, lesion / non-lesion | SUVmax | SUVmax / liver mean | Host-standardized SUVmax | Liver-referenced, host-standardized | Marginal normative (age, sex) | Conditional, linear | Conditional, linear + aleatoric SD | ΔAUC, conditional linear vs liver-referenced (95% CI) |
|---|---|---|---|---|---|---|---|---|---|---|
| PERCIST threshold | All scored hosts | 3,154 / 6,209 | 0.807 | 0.814 | 0.864 | 0.859 | 0.862 | 0.868 | 0.868 | 0.009 (−0.001 to 0.019) |
| | Organ hosts | 1,560 / 4,784 | 0.810 | 0.817 | 0.933 | 0.928 | 0.932 | 0.932 | 0.929 | 0.004 (−0.008 to 0.015) |
| | Region hosts | 1,594 / 1,425 | 0.755 | 0.759 | 0.780 | 0.775 | 0.781 | 0.790 | 0.791 | 0.015 (−0.000 to 0.029) |
| SUV 2.5 | All scored hosts | 2,502 / 26,065 | 0.960 | 0.951 | 0.906 | 0.907 | 0.907 | 0.916 | 0.917 | 0.009 (0.001 to 0.017) |
| | Organ hosts | 1,269 / 21,441 | 0.962 | 0.951 | 0.956 | 0.953 | 0.957 | 0.957 | 0.956 | 0.004 (−0.005 to 0.013) |
| | Region hosts | 1,233 / 4,624 | 0.939 | 0.933 | 0.905 | 0.905 | 0.905 | 0.911 | 0.912 | 0.006 (−0.004 to 0.015) |

AUC for lesion versus non-lesion hotspots of patients with malignant lesions (Supplementary Methods). With the PERCIST threshold, hotspots touched 91.9% of the 5,382 manual lesion components of at least 0.5 ml, and 59.0% of hotspots had a scored host; with SUV 2.5 the values were 99.9% and 66.4%. Host-standardized scores use the out-of-fold expected log SUVmax of the host organ or region and its scale; SUVmax and SUVmax / liver mean are not standardized by host. AUC, area under the receiver operating characteristic curve; CI, confidence interval.

**Supplementary Figure S1.** Distribution of deviation scores in control organs. (a) Quantile–quantile plot of *z* in control organs against the standard normal distribution. (b) Share of control organs above one-sided thresholds; black bars, nominal rates.

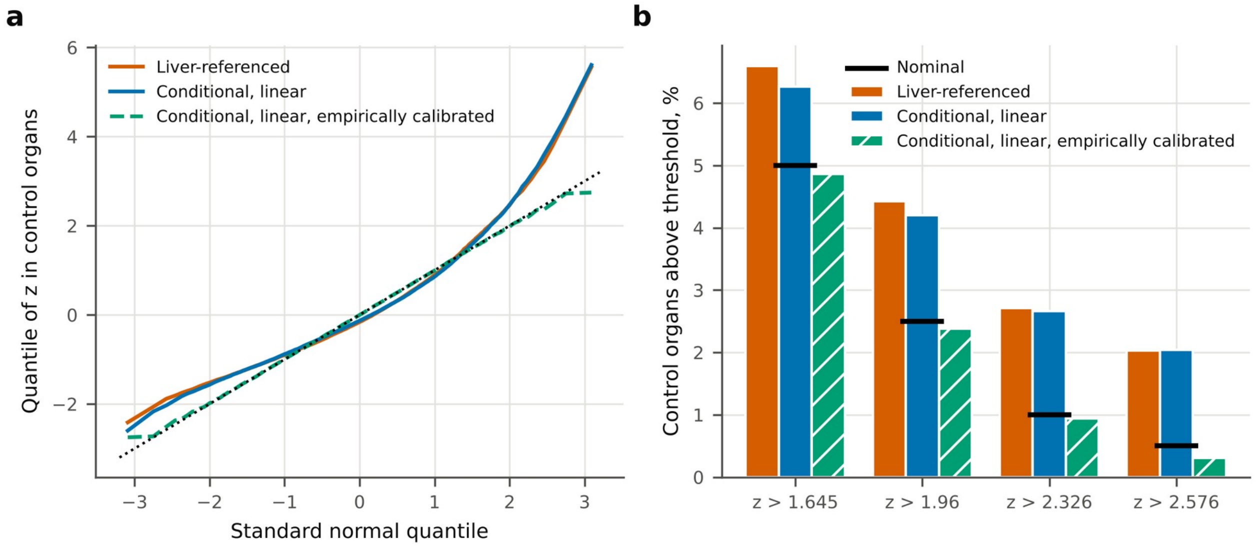


Liver-referenced and conditional linear scores are right-skewed because log SUVmax is a maximum; both exceed the nominal rates in the upper tail and fall short in the lower tail. Empirical calibration against the controls of the other folds follows the diagonal up to the largest control score, beyond which scores are tied.
