## Supplementary material for "Conditional normative modeling of whole-body FDG PET/CT: inter-organ relationships outperform reference-region normalization for label-free detection of lesion-bearing organs": CLAIM 2024 checklist

### Checklist for Artificial Intelligence in Medical Imaging (CLAIM): 2024 Update

| Section / Topic | No. | Item | Page / Line | No | NA |
|---|---|---|---|---|---|
| TITLE / ABSTRACT | | | | | |
| | **1** | Identification as a study of AI methodology, specifying the category of technology used (e.g., deep learning) | p. 1, l. 1–3; p. 2, l. 25–28; pp. 5–6, l. 136–146 | | |
| ABSTRACT | | | | | |
| | **2** | Summary of study design, methods, results, and conclusions | p. 2, l. 18–48 | | |
| INTRODUCTION | | | | | |
| | **3** | Scientific and/or clinical background, including the intended use and role of the AI approach | pp. 3–4, l. 51–84; p. 29, l. 755–759 | | |
| | **4** | Study aims, objectives, and hypotheses | p. 4, l. 85–91 | | |
| METHODS | | | | | |
| *Study Design* | **5** | Prospective or retrospective study | p. 30, l. 777–784 [a] | | |
| | **6** | Study goal | p. 4, l. 85–91; p. 8, l. 207–208 | | |
| *Data* | **7** | Data sources | p. 4, l. 94–102; p. 30, l. 770–773 | | |
| | **8** | Inclusion and exclusion criteria | p. 4, l. 102–103; p. 5, l. 116–119; p. 13, l. 345–352 | | |
| | **9** | Data pre-processing | pp. 4–5, l. 105–121; pp. 10–11, l. 269–283; p. 11, l. 285–288 | | |
| | **10** | Selection of data subsets | p. 4, l. 102–103; p. 11, l. 290–302; pp. 11–12, l. 305–314 | | |
| | **11** | De-identification methods | p. 30, l. 777–784 | | |
| | **12** | How missing data were handled | p. 5, l. 112–113; p. 5, l. 119–121; p. 10, l. 278–279; p. 13, l. 351 | | |
| | **13** | Image acquisition protocol | p. 4, l. 96; p. 28, l. 722–725 [b] | | |
| *Reference Standard* | **14** | Definition of method(s) used to obtain reference standard | p. 4, l. 98–100; p. 8, l. 208–213 | | |
| | **15** | Rationale for choosing the reference standard | p. 4, l. 98–100; p. 8, l. 208–213 | | |
| | **16** | Source of reference standard annotations | p. 4, l. 98–100 [c] | | |
| | **17** | Annotation of test set | p. 4, l. 98–100; p. | | |

| | | | | | |
|---|---|---|---|---|---|
| | | | 7, l. 175–178 | | |
| | **18** | Measures of inter- and intra-rater variability of features described by the annotators | [d] | | **X** |
| *Data Partitions* | **19** | How data were assigned to partitions | p. 7, l. 175–178; p. 11, l. 290–292 | | |
| | **20** | Level at which partitions are disjoint | p. 7, l. 175; p. 11, l. 290–292 | | |
| *Testing Data* | **21** | Intended sample size | [e] | **X** | |

| Section / Topic | No. | Item | Page / Line | No | NA |
|---|---|---|---|---|---|
| *Model* | **22** | Detailed description of model | pp. 5–6, l. 136–146; pp. 6–7, l. 148–192 | | |
| | **23** | Software libraries, frameworks, and packages | p. 4, l. 105; p. 12, l. 339–342 | | |
| | **24** | Initialization of model parameters | [f] | | X |
| *Training* | **25** | Details of training approach | pp. 5–6, l. 141–143; p. 7, l. 175–178; p. 7, l. 183–189 [g] | | |
| | **26** | Method of selecting the final model | pp. 5–6, l. 141–143; p. 6, l. 163–165 [h] | | |
| | **27** | Ensembling techniques | p. 7, l. 190–192 | | |
| *Evaluation* | **28** | Metrics of model performance | pp. 8–9, l. 213–231 | | |
| | **29** | Statistical measures of significance and uncertainty | p. 9, l. 217–218; p. 9, l. 238–239; p. 10, l. 260–261; pp. 11–12, l. 305–314 | | |
| | **30** | Robustness or sensitivity analysis | p. 9, l. 231–239; pp. 9–10, l. 241–256; p. 11, l. 299–302; p. 19, l. 464–476 | | |
| | **31** | Methods for explainability or interpretability | p. 6, l. 150–152; p. 8, l. 194–204; p. 8, l. 214–215 | | |
| | **32** | Evaluation on internal data | p. 7, l. 175–178 | | |
| | **33** | Testing on external data | [i] | X | |
| | **34** | Clinical trial registration | [j] | | X |
| RESULTS | | | | | |
| *Data* | **35** | Numbers of patients or examinations included and excluded | p. 13, l. 345–353; p. 22, l. 537–539 | | |
| | **36** | Demographic and clinical characteristics of cases in each partition | p. 13, l. 345–347; p. 13, l. 353 [k] | | |
| *Model performance* | **37** | Performance metrics and measures of statistical uncertainty | pp. 14–15, l. 378–393; p. 17, l. 425–441; p. 21, l. 511–513; p. 22, l. 539–544; p. 23, l. 572–578 | | |
| | **38** | Estimates of diagnostic performance and their precision | pp. 14–15, l. 378–393 | | |
| | **39** | Failure analysis of incorrect results | p. 18, l. 454–456; p. 19, l. 470–475; p. 21, l. 517–520; | | |

| | | | |
|---|---|---|---|
| | | | pp. 28–29, l. 735–738 |
| DISCUSSION | | | |
| | **40** | Study limitations | pp. 28–29, l. 722–753 |
| | **41** | Implications for practice, including intended use and/or clinical role | p. 29, l. 755–759 |
| OTHER INFORMATION | | | |
| | **42** | Provide a reference to the full study protocol or to additional technical details | p. 30, l. 774–775 [l] |
| | **43** | Statement about the availability of software, trained model, and/or data | p. 30, l. 770–775 |
| | **44** | Sources of funding and other support; role of funders | p. 30, l. 785–788 |

* Indicate page and/or line number for each checklist item that is present. NA = not applicable.

**Manuscript:** Conditional normative modeling of whole-body FDG PET/CT: inter-organ relationships outperform reference-region normalization for label-free detection of lesion-bearing organs. Page and line numbers refer to the submitted manuscript file with continuous line numbering.

**Notes**

[a] Retrospective secondary analysis of a public dataset (Ethics statement).

[b] Single-center, single-scanner data; acquisition and reconstruction protocols are described in the data descriptor [17].

[c] Lesion masks are the manual segmentations created and released by the dataset curators [17]; annotator details are given there.

[d] Lesion masks were used as distributed; no repeated or independent annotation was performed in this study.

[e] No a priori sample size calculation; all eligible studies were used (p. 4, l. 102–103). Dependence on training-set size was assessed empirically (p. 11, l. 299–302; Figure 6).

[f] Ridge regression and gradient-boosting models were fitted from scratch on tabular features; TotalSegmentator was used with its released pretrained weights (p. 4, l. 105).

[g] Five-fold patient-level cross-fitting with an inner five-fold split for score scaling and the variance model.

[h] Gradient-boosting settings were fixed before evaluation and ridge penalties were selected within the training data only; no model was selected on test-fold performance.

[i] No external test set; stated as a limitation (p. 28, l. 722–725).

[j] Not a clinical trial.

[k] Characteristics are given by diagnostic group and in total; folds were random patient-level splits of the whole cohort.

[l] No separate study protocol was registered; full analysis code is publicly available.
